# Graph Attention Networks for Drug Combination Discovery: Targeting Pancreatic Cancer Genes with RAIN Protocol

**DOI:** 10.1101/2024.02.18.24302988

**Authors:** Elham Parichehreh, Ali A. Kiaei, Mahnaz Boush, Danial Safaei, Reza Bahadori, Nader Salari, Masoud Mohammadi, Alireza Khorram

## Abstract

**Background:** Malignant neoplasm of the pancreas (MNP), a highly lethal illness with bleak outlook and few therapeutic avenues, entails numerous cellular transformations. These include irregular proliferation of ductal cells, activation of stellate cells, initiation of epithelial-to-mesenchymal transition, and changes in cell shape, movement, and attachment. Discovering potent drug cocktails capable of addressing the genetic and protein factors underlying pancreatic cancer’s development is formidable due to the disease’s intricate and varied nature.

**Method:** In this study, we introduce a fresh model utilizing Graph Attention Networks (GATs) to pinpoint potential drug pairings with synergistic effects for MNP, following the RAIN protocol. This protocol comprises three primary stages: Initially, employing Graph Neural Network (GNN) to suggest drug combinations for disease management by acquiring embedding vectors of drugs and proteins from a diverse knowledge graph encompassing various biomedical data types, such as drug-protein interactions, gene expression, and drug-target interactions. Subsequently, leveraging natural language processing to gather pertinent articles from clinical trials incorporating the previously recommended drugs. Finally, conducting network meta-analysis to assess the relative effectiveness of these drug combinations.

**Result:** We implemented our approach on a network dataset featuring drugs and genes as nodes, connected by edges representing their respective p-values. Our GAT model identified Gemcitabine, Pancrelipase Amylase, and Octreotide as the optimal drug combination for targeting the human genes/proteins associated with this cancer. Subsequent scrutiny of clinical trials and literature confirmed the validity of our findings. Additionally, network meta-analysis confirmed the efficacy of these medications concerning the pertinent genes.

**Conclusion:** By employing GAT within the RAIN protocol, our approach represents a novel and efficient method for recommending prominent drug combinations to target proteins/genes associated with pancreatic cancer. This technique has the potential to aid healthcare professionals and researchers in identifying optimal treatments for patients while also unveiling underlying disease mechanisms.

**Highlights:**

- Graph Attention Networks (GATs) used to recommend drug combinations for pancreatic cancer
- RAIN protocol applied to extract relevant information from clinical trials and literature
- Gemcitabine, Pancrelipase Amylase, and Octreotide identified as optimal drug combination
- Network meta-analysis confirmed the effectiveness of the drug combination on gene targets
- Novel and efficient method for drug discovery and disease mechanism elucidation

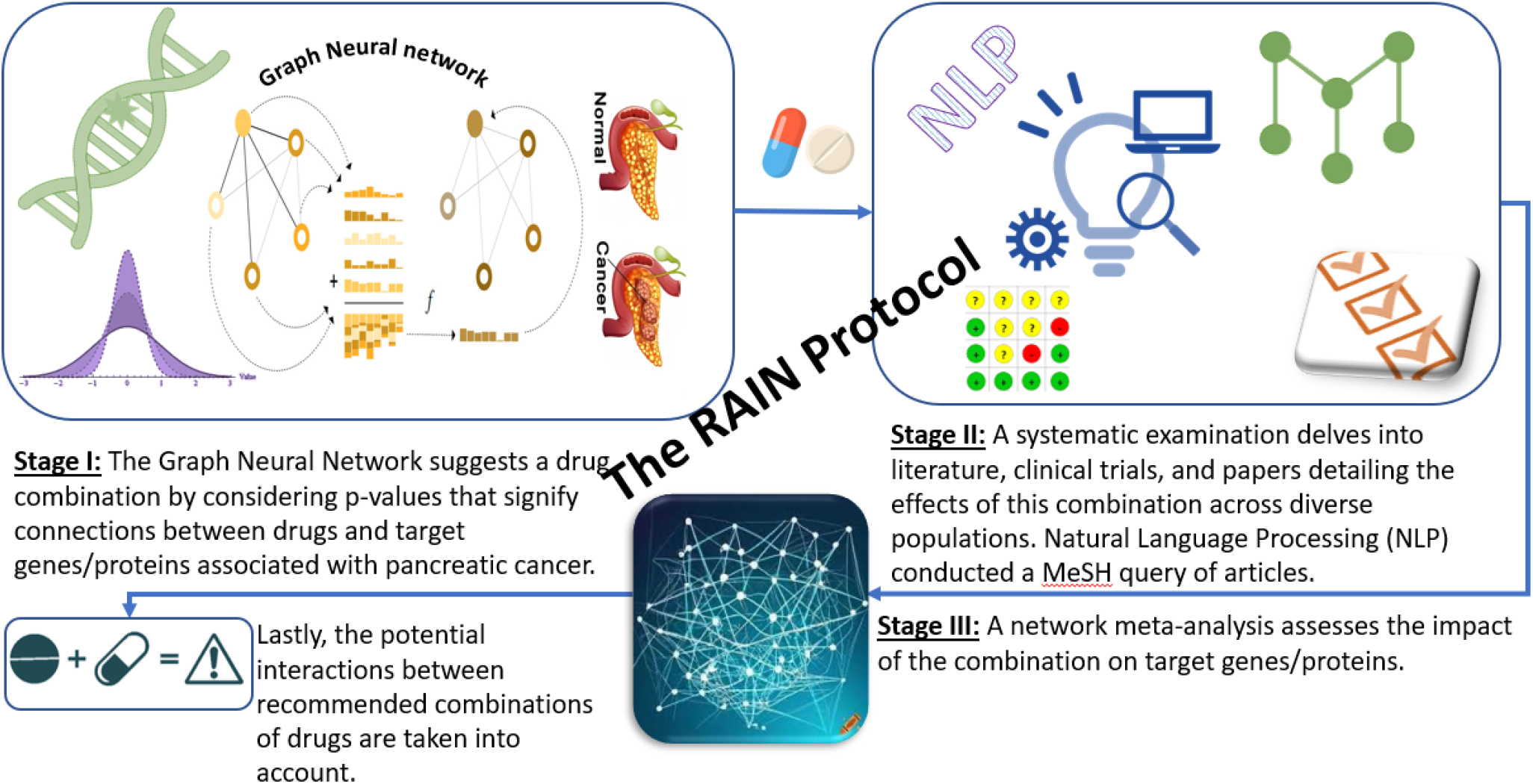

## 1. INTRODUCTION

Pancreatic cancer, formally known as a malignant neoplasm of the pancreas, is a severe condition marked by the unrestricted proliferation of irregular cells in the pancreas, leading to the creation of a tumor. Its prognosis is often unfavorable due to late-stage diagnosis and resistance to treatment. However, recent advancements in medical research and treatment techniques are positively impacting the outlook for individuals with pancreatic cancer. Enhanced diagnostic methods, such as comprehensive imaging and molecular testing at the time of diagnosis, aid in enhancing comprehension of cancer’s behavior and response to treatment. Accurate staging using PET scans and genetic testing is now employed. Mayo Clinic has introduced innovative genetic testing, analyzing patients’ blood and abdominal fluid through laparoscopy to detect cancer DNA, helping identify those at risk for recurrence and tailoring treatment for prevention. A study focused on pancreatic mucinous cystadenocarcinoma (MCAC) investigated clinical features’ prognostic value pertaining to the survival specific to cancer and the spread of cancer cells. A graphical tool was created to forecast individual results, guiding personalized clinical decisions. Researchers, including Park and colleagues, propose inhibiting a major inflammatory pathway to sensitize pancreatic tumors to chemotherapy and immunotherapy. Advances in treatment are enhancing patient outcomes, enabling long-term survival. Managing pancreatic cancer is crucial, with neoadjuvant therapy proving effective in altering the immune microenvironment, improving resection rates, eliminating micro metastases, transforming unresectable tumors into candidates for surgery, and reducing recurrence. [1], [2], [3]

Research has been conducted to examine the predictive significance of clinical characteristics in predicting cancer-specific survival (CSS) and metastasis in individuals with a diagnosis of pancreatic mucinous cystadenocarcinoma (MCAC). The objective of these studies is to create a reliable nomogram for predicting CSS. Factors such as age exceeding 65 years, poorly differentiated or undifferentiated tumors, and primary tumor resection emerged as independent risk factors influencing overall survival time. A healthcare professional, particularly a physician or endoscopist, should address aspects like pain management, nutritional concerns (including endocrine and exocrine insufficiency), biliary obstruction, and thromboembolic risk. Ongoing research aims to deepen the understanding of pancreatic cancer to develop more efficient treatment strategies. In summary, effective management of pancreatic malignant neoplasm plays a vital role in improving patient survival, reducing recurrence, and enhancing overall quality of life.[4], [5], [6]

### 1.1. Associated human genes/Proteins

Mutations in the KRAS gene are some of the most common genetic changes. observed in cancer, with a notable occurrence in pancreatic cancer. The KRAS G12D mutation stands out as the most frequently encountered KRAS mutation, prevalent in the majority of pancreatic tumors with KRAS mutations. Despite being historically considered “undruggable,” recent breakthroughs have provided optimism in targeting KRAS G12D. A noteworthy development involves MRTX1133, a compound designed to attach to and hinder KRAS G12D. Another promising approach involves immunotherapy, utilizing utilize adoptive T-cell transfer to selectively address G12D in pancreatic cancer. Positive outcomes were observed in a patient with metastatic G12D-mutated pancreatic cancer, showing regression of metastases following this therapy. These advancements suggest that KRAS, especially the G12D mutation, holds potential as a focal point for addressing pancreatic malignant neoplasms in therapy.[7], [8]

The somatostatin receptor (SST) is widely distributed in tumors from various organs and presents a promising target for Theranostic applications. While current therapies focusing on SST are primarily limited to typical gastroenteropancreatic neuroendocrine tumors (NETs), there is potential to expand the scope of these treatments. Subtype 2 of somatostatin receptor (SST2) is the primary focus of peptide receptor radiotherapy (PRRT). A comprehensive analysis across different cancer types revealed diverse SST2 expression levels. Notably, low-grade glioma (LGG) and breast invasive carcinoma (BRCA) exhibited elevated SST2 expression. The level of The expression of SST2 exhibited noteworthy results. associations with genomic and clinical factors across various cancers. These findings propose that SST, particularly SST2, could serve as a potential target for the treatment of pancreatic malignant neoplasms. Yet, additional investigation and clinical trials are needed. are essential to fully comprehend the capability of these therapeutic approaches.[9]

Inherited as an autosomal dominant condition is Multiple Endocrine Neoplasia Type 1 (MEN1). with autosomal dominant inheritance. arising from genetic alterations in the tumor suppressor gene MEN1. This gene codes for menin, a 610-amino acid protein. Mutations in MEN1 can disrupt signaling pathways, leading to systemic endocrine disorders, such as pancreatic neuroendocrine tumors (pNETs). MEN1 stands as the predominant cause of inherited PanNETs. The development of these diseases can be attributed to the loss of menin expression or abnormal nuclear translocation induced by mutations in the MEN1 gene.[10], [11], [12]

SMAD4, also recognized as the DPC4 gene (Deleted in Pancreatic Cancer), belongs to an extensive family of proteins situated located on chromosome 18q and functions as the main transducer of signals for the TGF-β family. It stands out as a major tumor-suppressive gene specifically targeted in infiltrating pancreatic cancer (PC), with its inactivation being relatively specific to this type of tumor. Notably, SMAD4 mutations are now acknowledged in over 50% of pancreatic ductal carcinomas. SMAD4 has a vital function in the signaling pathway of transforming growth factor-β (TGF-β). Initially, TGF-β acts as a tumor suppressor in pancreatic tumorigenesis via the phosphorylation and stimulation of the SMAD4/DPC4 gene. However, the mutation in the SMAD4/DPC4 gene triggers the development of cancer and the advancement of tumors in different types of cancers, such as lung, colon, and pancreatic cancer. A SMAD4-negative status is associated with a higher propensity for metastasis rather than local recurrence. A specific genetic variation of pancreatic ductal adenocarcinoma (PDAC), identified by the loss of both alleles of DPC4 in addition to a missense mutation in TP53, is associated with heightened metastatic effectiveness. Additionally, CHGA is commonly utilized as a marker for neuroendocrine tumors, including pancreatic neuroendocrine tumors. [13], [14], [15], [16]

Mesothelin (MSLN) is a glycoprotein found in different tumors, such as mesothelioma, ovarian cancer, and pancreatic cancer, with varying expression levels, and others. In the past decade, there has been an increasing interest in regarding mesothelin (MSLN) as a potential antigen linked to pancreatic ductal adenocarcinoma (PDAC). The limited presence of MSLN in normal tissues such as peritoneum, pleura, and pericardium, coupled with its overexpression in 80 to 90% of PDAC cases, positions it as an appealing potential for therapeutic interventions in patients with pancreatic ductal adenocarcinoma (PDAC). Various MSLN-targeting agents have been discussed in selected articles, including tracers like The application of 64Cu-DOTA-11-25mAb anti MSLN, 111In-MORAb-009-CHX-A″, 89Zr-MMOT0530A, 111In-amatuximab, 99mTc-A1, 89Zr-AMA, 89Zr-amatuximab, 64Cu-amatuximab, 89Zr-labeled MMOT0530A, and 89Zr-B3 has been demonstrated in the detection of malignancies that show overexpression of mesothelin. [17] [18], [19]

PALB2 (Partner and Localizer of BRCA2) is a gene that, when mutated, elevates the risk of certain cancers, including pancreatic cancer. In a small subset of patients with pancreatic ductal adenocarcinoma (PDAC) who have mutations resulting in the loss of PALB2 function, there can be substantial differences in both initial and ongoing treatment approaches. These mutations lead to the loss of homologous recombination (HR) in double-strand break DNA repair, significantly impacting drug sensitivities. Poly (ADP-ribose) polymerase (PARP) inhibitors like Niraparib, Olaparib, Talazoparib, Rucaparib, and Veliparib are now approved for various cancers with impaired high-fidelity double-strand break HR, including those with deleterious mutations in BRCA1/2, PALB2, and functionally related genes. Recent findings indicate that the presence of such mutations in PDAC notably influences responses to drugs, both in terms of initial chemotherapy and ongoing maintenance therapy.[20]

The CDKN2A gene, also known as Cyclin Dependent Kinase Inhibitor 2A, plays a vital role as a tumor suppressor by coding for two essential proteins, namely p16 INK4A and p14 ARF that have a crucial function in controlling pathways related to the cell cycl. Both genetic and epigenetic alterations that deactivate CDKN2A are commonly observed in various cancers, including pancreatic cancer. Germline changes in CDKN2A are most frequently linked to a predisposition for melanoma and pancreatic cancer. Tumor tissue tends to exhibit higher CDKN2A expression compared to normal tissue, serving as a prognostic indicator for patients with tumors. The expression level of CDKN2A has significant correlations with tumor mutation burden (TMB) in 10 different cancers, and it is also associated with microsatellite instability (MSI) in the same set of tumors. Furthermore, the expression of CDKN2A is associated with levels of infiltrating lymphocytes (TIL). across 22 different cancers, suggesting an association with tumor immunity. BRCA2(Breast Cancer susceptibility gene 2) is another gene, and its mutations are associated with an elevated risk of certain cancers, including pancreatic cancer. Research studies have established a connection between BRCA2 gene mutations and pancreatic cancer, indicating that individuals testing positive for a BRCA2 mutation have a heightened likelihood of developing pancreatic cancer. In cancers with a BRCA2 mutation, a unique vulnerability is created that can be targeted by PARP inhibitors. These inhibitors hinder repairing single-strand DNA breaks, causing them to transform into double-strand breaks. According to NCCN guidelines, individuals with metastatic or regionally advanced pancreatic cancer carrying deleterious germline BRCA1/2 or PALB2 mutations are advised to commence platinum-based chemotherapy as their primary therapy. The recommendations also propose the potential use of Olaparib for ongoing treatment in individuals with harmful germline BRCA1/2 mutations, guided by the findings from the phase 3 POLO trial.[21] [22], [23]

TP53 is a crucial gene that inhibits tumor formation, and alterations in this gene are detected in more than half of all human cancers. These genetic changes not only hinder the gene’s While diminishing antitumor effectiveness, these mutations also bestow oncogenic characteristics. to the mutant p53 protein. The method for p53-targeted therapy originated from detecting substances with the ability to revive or rekindling the functions of wild-type p53 or eliminating mutated p53.Focusing on mutated p53 are highly dependent on the structure and specific drug species. The alteration of the wild-type p53 disrupts various pathways for survival typically sustained by it, leading to the initiation of alternative genes or pathways that support the survival of cancer cells. Moreover, as the cancer-promoting functions of mutated p53 contribute to the proliferation and spread of cancer, focusing on the pathways of signaling affected by p53 mutation appears to be an attractive approach. Synthetic lethality refers to a situation where disruption of one of the genes alone is tolerable in two genes displaying interactions leading to synthetic lethality, but total inactivation of both genes leads to cellular demise. Hence, instead of specifically aiming at p53, exploiting genes with synthetic lethal interactions with mutant p53 may offer additional therapeutic advantages. [24]

MUC1 (Mucin 1) is a glycoprotein with diverse roles in maintaining normal bodily functions and contributing to the development of cancer. It is notably elevated in various cancers, with pancreatic cancer showing predominant overexpression. MUC1 has a part in inhibiting interactions between cells and between cells and the surrounding tissue. It also acts as a signal transducer, actively participating in the advancement of cancer. Indications propose that MUC1 serves as a marker for an aggressive cancer characteristic and could promote the dissemination of cancer cells through blood vessels. Recently, a therapeutic effect against a pancreatic cancer model was observed with a monoclonal antibody-drug conjugate that targets MUC1. This antibody attaches to the surface of pancreatic cancer cells where MUC1 is present, effectively restraining inhibiting cell growth through the induction of G2/M cell cycle arrest and apoptosis. Significantly, it demonstrated a substantial reduction in suppressing the development of pancreatic xenograft tumors by hindering cell proliferation and encouraging cell death.[25]

MUC4, identified as a transmembrane mucin, is involved in development of Pancreatic cancer is present in various normal and malignant tissues. Its functions in the renewal and differentiation of epithelial tissue are noteworthy. Recent research indicates that MUC4 may have opposing effects on prognosis, proliferation, metastasis, and immune response. It has been linked to genomic alterations, tumor proliferation, metastasis, and tumor infiltration. These discoveries underscore the significance of a comprehensive evaluation. of MUC4 as both a biomarker and a potential therapeutic target. Additionally, MUC4 influences HER2/ErbB2 signaling and significantly impacts the therapeutic outcomes of Herceptin-based therapy, underscoring its potential utility in cancer treatment and planning.[25] [26] [27], [28]

PRSS1, also known as the cationic trypsinogen gene, is associated with chronic pancreatitis and the development of pancreatic cancer. Somatic mutations in PRSS1 have been identified in individuals with these conditions. However, the precise role of PRSS1 mutations in triggering the mechanisms behind either fostering malignant proliferation and metastasis in pancreatic cancer or promoting these aspects are not yet fully understood. One study proposed that mutation and excessive expression of PRSS1 might contribute to an “inside job” mechanism in the formation of pancreatic cancer and the growth of tumors. The study demonstrated a significant increase in migration and infiltration in PANC-1 cells harboring the R116C mutation compared to their in a transgenic mouse model expressing iZEG-PRSS1_R116C, primary pancreatic intraepithelial neoplasia (PanINs) was detected within the pancreatic duct, indicating a novel pathway involved in facilitating the development of pancreatic cancer, distinct from its wild-type counterparts. However, these areas of research are ongoing, and the exact mechanisms are still under exploration. For more specific and detailed information, it is advisable to consult with a healthcare professional or a researcher in this field.[29]

Glucagon (GCG) is a hormone synthesized by alpha cells in the pancreas, playing a vital role in regulating glucose levels by promoting hepatic glucose production. However, the specific involvement of GCG in pancreatic neoplasms is not thoroughly investigated, and the existing literature lacks a comprehensive mechanism explaining how GCG could serve as a potential target for malignant pancreatic neoplasms^4^. Further research is necessary to fully comprehend GCG’s role in the development and treatment of pancreatic neoplasms. It’s noteworthy that progress in understanding the immune microenvironment within the tumor (TIME) and the development of therapies focusing on TIME components show promise in treating pancreatic ductal adenocarcinoma (PDAC), an extremely deadly solid tumor.[30] [3]

SLC29A1, also recognized as equilibrative nucleoside transporter 1 (ENT1), is involved in the transportation and resilience associated with the nucleoside analog cytosine arabinoside (AraC), a highly effective drug in treating acute myeloid leukemia (AML). Nevertheless, the particular role of SLC29A1 in pancreatic neoplasms remains inadequately explored, and the existing literature lacks a comprehensive mechanism. Further investigation is required to gain a complete understanding of SLC29A1’s role in the development and treatment of pancreatic neoplasms.[31] [32]

CDH1, also identified as E-cadherin, is a gene responsible for encoding a a protein integrated into the plasma membrane of epithelial cells, creating tissues that coat the surfaces of the body and line them different hollow spaces, passages, and glands. It is essential for preserving the integrity of epithelial tissues, and the absence of E-cadherin has traditionally been regarded as a characteristic feature of metastatic cancers, providing cancer cells capable of migration and invade neighboring tissues. However, recent in-depth analysis of various Examination of databases indicates that CDH1 mRNA and E-cadherin protein are not uniformly reduced in the majority of carcinoma tissues and cell lines. Contrary to the conventional understanding of E-cadherin loss Throughout tumor advancement and the spread of cancer, the levels of CDH1 mRNA and E-cadherin protein either increase or stay constant in numerous carcinoma cells when compared to normal cells. Additionally, CDH1 mRNA levels show a positive correlation with the survival of cancer patients. These findings suggest that the role of E-cadherin the explanation for tumor advancement and metastasis might have been overly simplified in the past. The levels of CDH1 mRNA could function as a dependable biomarker for diagnosing certain cancers (like colon and endometrial carcinomas) because of the notable increase in CDH1 mRNA during the initial phases of tumor formation in these carcinomas.[33]

Somatostatin Receptor Type 2 (SSTR2) is a protein widely distributed in various tissues and extensively expressed in solid tumors, positioning it as a prospective target for therapeutic intervention for cancer, especially neuroendocrine tumors (NETs), because of its heightened expression in these malignancies. Concerning neuroendocrine tumors in the pancreas (PNETs), SSTR2 plays a significant role. A study demonstrated that the enhancement of SSTR2 in an pancreatic neuroendocrine tumor model within a living organism deficient in receptors improved the tumor’s response to targeted 177Lu-DOTATATE therapy^2^. This therapy involves the use of a radiolabeled somatostatin analog binding to SSTR2, enabling precise radiation therapy. Additionally, SSTR2 has been identified as highly methylated in colorectal cancer and involved in the carcinogenesis of gastric and Breast cancers are influenced by the interaction of somatostatin with SSTR2, which also inhibits the release of cytokines from immune cells, thereby affecting the tumor microenvironment. (TME).[34] [35]

TSC1, also referred to as Tuberous Sclerosis Complex, is a crucial component of the pro-survival PI3K/AKT/mTOR signaling pathway. It has a crucial function in fundamental processes like development, cell growth, proliferation, survival, autophagy, and cilia development, working in conjunction with different regulatory molecules. Current investigations has underscored the inhibitory effect on tumors TSC1’s role in numerous human cancers, such as those affecting the liver, lung, bladder, breast, ovarian, and pancreas, has been identified. TSC1 integrates inputs from a variety of sources communication pathways, thereby regulating cancer cell activities like proliferation, metabolism, migration, invasion, and immune regulation. Regarding pancreatic cancer, the removal Activation of Tsc1 in the pancreatic context initiates mTORC1 signaling, resulting in noticeable lesions resembling adenocarcinoma with metastasis to the liver and lungs.[36]

Serine Protease Inhibitor Kazal Type 1 (SPINK1), belonging to the SPINK family, is significantly involved in the normal functioning of the pancreas. and is implicated in pancreatic neoplasms. Its expression is closely linked to human tumors and is induced under hypoxic conditions commonly found in tumor microenvironments, resulting in elevated levels of secreted SPINK1^2^. Both xenografted and clinical tumor tissues exhibit the presence of SPINK1 proteins within and around hypoxic regions. Notably, these secreted SPINK1 proteins contribute to enhancing the. The resistance of cancer cells to radiotherapy, even in the face of radiation in Normal oxygen levels. Regarding radiotherapy, SPINK1 emerges as a possible focus for therapeutic intervention radiosensitization. The application of a neutralizing antibody targeting SPINK1 has demonstrated a enhancement of sensitivity to radiation. These results imply that SPINK1, released by cells experiencing low oxygen levels, acts in a paracrine manner to shield surrounding cancer cells with sufficient oxygen levels from radiation, highlighting its role in radioresistance.[37] [38]

FOXM1 (Fork head Box Protein M1) is a crucial transcription factor linked to cellular proliferation, self-renewal, and tumorigenesis. Its widespread expression during the cell cycle links it to cancer, where its overexpression is commonly observed, signifying an unfavorable outlook for individuals with cancer. FOXM1 has a crucial function in maintaining cancer hallmarks by regulating target regulation of gene expression at the transcriptional stage. Designated as the 2010 Molecule of the year for its capacity as a molecular focal point in the treatment of cancer, the exact cause of FOXM1 irregularity is still unclearA thorough comprehension of the control mechanisms governing FOXM1 holds promise for insights into various diseases where FOXM1 is pivotal. This summary covers the control of FOXM1 at the transcriptional, post-transcriptional, and post-translational levels, providing crucial significance for developing innovative approaches aimed at FOXM1. In the realm of pancreatic neoplasms, ongoing investigations seek to unravel the specific mechanisms by which FOXM1 acts as a potential target. Notably, disrupting the Site where FOXM1 binds inhibits Verification of the activity of the FOXM1 promoter significance of The auto-regulatory activation of the −745/−738 bp region. Intriguingly, FOXM1 peptides Induce cytotoxic T lymphocytes (CTLs) specific to HLA-A2 in transgenic mice expressing HLA-A2 indicating FOXM1’s potential suitability as a target for immunotherapy against cancers.[39]

MUC2, a mucin type, is a glycoprotein with diverse functions in maintaining balance and contributing to carcinogenesis. Within the context of pancreatic neoplasia, MUC2 serves not only as an indicator of an indolent pathway but also contributes to the less aggressive nature of such tumors. Aggressive pancreatic tumors typically exhibit rare detectability of MUC2, while it is frequently present in intraductal papillary mucinous neoplasms (IPMNs) are rare and known for their slow-growing characteristics.Therefore, in the realm of pancreatic neoplasia, MUC2 holds possible diagnostic and prognostic significance a marker for indolent phenotypes.[25]

Ring Finger Protein 43 (RNF43), functioning as an E3 ubiquitin ligase, has been identified with mutations in several cancers. In the context of pancreatic neoplasms, RNF43 emerges as an indicator for the the malignant conversion of cystic mucinous lesions in the pancreas neoplasms (MCNs). The presence of RNF43 was notably decreased in 71% of cases with high-grade dysplasia or invasion carcinoma (HG/INV) cases, showing significant correlation with histological grade and aberrant expression of β-catenin. Moreover, there was a notably elevated occurrence of RNF43 mutations in cases with high-grade dysplasia or invasion compared to low-grade dysplasia (LG) cases. These results imply that mutations in RNF43 might be involved in and serve as predictors of malignant transition from the initial phase of mucinous cystic neoplasm (MCN).However, the specific mechanisms through which RNF43 acts as a potential target for malignant neoplasms of the pancreas are still under investigation, requiring further research for a comprehensive understanding of its role in the pathogenesis and treatment of pancreatic neoplasms.[40] [41]

Vimentin (VIM) is a type of intermediate filament (IF) protein, often upregulated during transition from epithelial to mesenchymal state (EMT) and in the context of cancer, particularly during invasion and metastasis. However, the specific involvement of VIM in pancreatic neoplasms lacks thorough investigation, with the existing literature lacking a detailed mechanism. Further research is necessary to gain a comprehensive understanding of VIM’s role in the pathogenesis and treatment of pancreatic neoplasms. It’s worth noting that advancements in comprehending the immune milieu within the tumor (TME) and the development of therapies targeting TIME components show promise in treating pancreatic ductal adenocarcinoma (PDAC), an extremely deadly solid tumor.[42]

Poly (ADP-ribose) polymerase 1 (PARP1) is a crucial enzyme engaged in the repair of DNA, and it has been identified as a possible target for cancer. therapy, particularly when combined with immune checkpoint inhibitors (ICIs). PARP1 inhibitors (PARPi) represent a category of medications that hinder the repair of single-strand DNA, causing DNA damage and triggering apoptosis. Recent evidence suggests that PARPi may have the ability to influence stimulating the immune response against tumors by activating antigen-presenting cells, promoting the infiltration of effector lymphocytes, and increasing the presence of programmed death ligand-1 in tumors. This implies that targeting PARP1 could be a viable approach for treating malignant neoplasms of the pancreas, particularly when used in combination with ICIs. Nonetheless, additional research is required to acquire a comprehensive understanding of PARP1’s function in the pathogenesis and treatment of pancreatic neoplasms.[43]

GNAS is a proto-oncogene that holds significance in the case of intraductal papillary mucinous neoplasm (IPMN) occurring in the pancreas. Mutations specific to IPMNs are identified in GNAS, occurring at a rate of 41–75%. These mutations are likely involved in the progression of IPMNs following the emergence of neoplastic cells, instead of in the initiation of IPMNs. Kawabata et al. have demonstrated that mutant GNAS restrains tumor aggressiveness in established pancreatic cancer by counteracting the pathway involving KRAS. Their findings indicate that grafts of cells with wild-type GNAS exhibit an elevated Ki-67 labeling index compared to GNAS-mutant cells. Notably, GNAS wild-type tumors undergo phenotypic changes, leading to a notable decrease in the production of mucin and the presence of solid components with massive stromal elements. Transcriptional profiling suggests a clear discord between mutant GNAS and KRAS signaling.[44] [45] [46] [47]

S100P, belonging to the group of proteins known as the S100 family, is involved in different types of cancers and emerges as a possible indicator for the immune-suppressed environment in pancreatic cancer. Research by Hao et al. has identified differential presence of S100P in pancreatic cancer, associating it with a poor prognosis. Significantly, S100P exhibits a notable inverse relationship with the infiltration of immune cells, especially CD8+ T cells. Additionally, a strong connection association between S100P and immunotherapy is recognized, given its significant correlates with tumor mutation burden (TMB) and the levels of expression for TIGIT, HAVCR2, CTLA4, and BTLA. Interestingly, elevated S100P levels demonstrates an inverse association with levels of methylation, linked to CD8+ T cells. In vitro RT-PCR confirms increased S100P expression in all five pancreatic cancer cell lines, and immunohistochemical (IHC) analysis confirms elevated levels of S100P in pancreatic cancer tissues. These results propose S100P as a potential indicator for the the environment suppressing the immune system, potentially offering a novel therapeutic avenue for pancreatic cancer. However, further research is essential to comprehensively understand the role of S100P in the pathogenesis and treatment of pancreatic neoplasms. DAXX (Death Domain-Associated Protein) is a protein identified as be mutated in several cancers. In the context of pancreatic neoplasms, particularly pancreatic neuroendocrine tumors of the pancreas (PanNETs), DAXX emerges as a predictor of malignant transformation. Wang’s study results reveal significantly shortened survival without disease recurrence and survival without relapse in individuals with modified DAXX genes, through combined hazard ratios (HRs) of 5.05 and 3.21, respectively. Yet, there is no distinction in the overall survival among individuals with modified DAXX genes or not, with a combined HR of 0.71. These findings indicate that mutations in DAXX might play a role in and serve as predictors of the progression to malignancy from an early stage of the PanNETs.[48], [49] [50]

GLI1 serves as a transcriptional effector in the Hedgehog signaling (Hh) pathway, carefully controlled in embryonic development and tissue pattern/differentiation. Aberrant activation of GLI1 in certain cancers is associated with promoting various cancer hallmarks, including growth, viability, blood vessel formation, spread to other tissues, metabolic alterations, and resistance to chemotherapy. In In pancreatic cancer, prevalent activating mutations occur in either KRAS or BRAF genes. lead to the activation of cancer-causing pathways such as RAS/RAF/MEK/ERK, PI3K-AKT-mTOR, and TGFβ, converging on GLI1 activation. This activation contributes to cell growth, advancement of tumors, resistance to chemotherapy, and early spread. Understanding the mechanisms underlying GLI1 dysregulation can offer predictive and diagnostic markers for identifying patients who would benefit therapeutically either by directly inhibiting GLI1 or by employing targeted therapy focused on proteins controlled downstream of GLI1.[51]

MUC5A, a mucin type characterized by heavy glycosylation and the formation of intricate polymers, plays a crucial role in constructing the framework of polymeric mucus gel on epithelial cell surfaces. Its functions are diverse, encompassing barrier functions to epithelial cells, interactions between hosts and pathogens, attraction of immune cells to premalignant or malignant lesion sites, and tumor progression in a context-dependent manner. Ongoing efforts involve the development of overexpression strategies and genetically engineered mouse models for the study of this structurally complex and evolutionarily conserved gel-forming mucin. MUC5A is increasingly recognized as a potential target for diagnosis, prognosis, and therapeutic interventions across various malignancies.[52]

MUC6, a mucin type characterized by heavy glycosylation and the formation of intricate polymers, plays a crucial role in constructing the structure of polymeric mucus gel on the surfaces of epithelial cells. Its functions are diverse, encompassing barrier functions to epithelial cells, interactions between hosts and pathogens, attraction of immune cells to potentially cancerous or malignant lesion sites, and the advancement of tumors in a manner dependent on the context. Ongoing efforts involve the development of overexpression strategies and mouse models engineered at a genetic level for the study of this intricate and evolutionarily mucin that forms a conserved gel. MUC6 is increasingly recognized as a potential focus for diagnosis, prognosis, and therapeutic interventions across various malignancies.[53]

PDX1 serves as a key controller in the development of the pancreas, overseeing regulator in the formation of the pancreas maintenance of β-cells, crucial for regular functioning of insulin^1^. Alterations in the PDX1 gene are associated experiencing different pancreatic abnormalities, ranging from absence of the pancreas (in cases of a mutation in both alleles) to MODY4 (in instances of a mutation in one of the two alleles). Conversely, reduced PDX1 presence is noted in various forms of diabetes. In the realm of pancreatic cancer, PDX1 has been recognized as a promising target for therapy. Studies indicate that PDX1 exhibits oncogenic properties in pancreatic cancer and may also function as an indicator in other solid human tumors, including those affecting the colon, prostate, and kidney.[54] [55]

BRCA1, a gene crucial for DNA repair, functions as a gene suppressing tumors, undergoing mutations impacting the likelihood of developing pancreatic cancer and influencing treatment decisions. Mutations in BRCA1 elevate the likelihood of various cancers, including pancreatic cancer. Individuals may carry one healthy and one mutated BRCA1 copy, leading to an elevated but not definite cancer risk. Cancer may arise if the second, healthy copy undergoes mutations due to environmental exposures or cellular genetic errors. A targeted cancer drug already utilized for certain ovarian and breast cancers may also benefit individuals with advanced pancreatic cancer harboring inherited BRCA1 or BRCA2 mutations, as per findings from a substantial clinical trial. The BIRC5 gene, accountable for producing the Survivin protein, belongs a member of the inhibitor of apoptosis family. While Survivin is typically present in the course of embryonic development and absent in cells in adulthood, it is prevalent in the majority of cancer cells, rendering it a hopeful target for anticancer medications and a possible prognostic indicator.A comprehensive cancer analysis identified thorough analysis of various cancers, revealing significantly higher expression in cancer tissues across 16 cancer types in comparison to healthy tissues. Higher BIRC5 expression associated with worse overall survival in 14 out of 33 cancer types. Although the specific role of BIRC5 in pancreatic cancer wasn’t detailed, controlled for age and tumor grade, BIRC5 the level of expression adversely affected overall survival in various cancers. This suggests a potential similar role in pancreatic cancer, but dedicated research is essential to confirm. Despite its anti-apoptotic nature promoting tumor progression, BIRC5 is considered a potential therapeutic target, supporting its potential relevance in addressing malignant neoplasms of the pancreas.. [56] [57]

ZEB1, a transcriptional repressor, holds plays a vital role in the progression of different epithelial tumors, including pancreatic cancer, by triggering the epithelial-mesenchymal transition (EMT). This contribution to the process migration, invasion, and the formation of metastasis in carcinoma cells. In pancreatic cancer, ZEB1 is recognized as an EMT inducer, leading to the loss of epithelial differentiation and the acquisition of a mesenchymal characteristic. This transition enables cancer cells to detach from the main tumor mass and spread into the surrounding stroma. The downregulation of E-cadherin expression, facilitated by ZEB1, often accompanies this process. Additionally, ZEB1 is linked to the acquisition of cancer stem cell traits, supporting the migrating cancer stem cell (MCSC) hypothesis. This implies that ZEB1 not only enhances the invasive capabilities of pancreatic cancer cells but also contributes to their resistance against treatments. In essence, ZEB1’s involvement in EMT, its interaction with molecules like E-cadherin and microRNAs, positions it as a potential focus for therapeutic interventions in malignant neoplasms of the pancreas. However, more dedicated research is essential to comprehend the mechanisms and explore potential therapeutic applications further. For detailed information, it is advisable to consult with a healthcare professional.[58] [59]

Epithelial cell adhesion molecule (EPCAM) is a glycoprotein with a homophilic type I transmembrane structure. known to play diverse roles in normal physiological conditions processes as well as in cancer development and progression. EPCAM is recognized for its involvement in cellular adhesion, acting not only serving as a ligand for transmitting signals receptors but also acting as a signaling ligand. These multifaceted molecular mechanisms contribute to the complexity of EPCAM’s role in cancer advancement. EPCAM’s engagement in cancer stem cell characteristics, cell growth, metabolic processes, angiogenesis, the transition from epithelial to mesenchymal states (EMT), the spread of cancer to other sites, resistance to chemotherapy and radiation, and the modulation of the immune system. has been identified. Throughout advancement of tumors, EPCAM engages in interaction with crucial signaling pathways like Wnt/β-catenin, TGF-β/SMAD, EpEX/EGFR, PI3K/AKT/mTOR, and p53, instigating alterations in the biology of cancer cells. Although specific studies in the context of pancreatic cancer were not uncovered in the search results, the roles of EpCAM in other cancers suggest its potential as a target for malignant neoplasms of the pancreas. Its participation in critical processes like EMT and metastasis, coupled with interactions with other molecular players, positions it as a viable target for therapeutic interventions.[60] [61]

The underlying mechanism mammalian the focal point of rapamycin (mTOR) is a pivotal component of the the PI3K-Akt-mTOR signaling cascade, exerting significant influence on diverse biological activities such as cellular proliferation, survival, metabolism, autophagy, and immunity. Anomalies in initiation of this signaling cascade may create a cellular environment conducive to transformation. In the realm of cancer, dysregulation of this system, characterized by genetic mutations and overexpression, has been associated with numerous cancers in humans. As a result, mTOR has become a vital focus for cancer therapy., particularly for cancers exhibiting heightened mTOR signaling as a result of genetic or metabolic irregularities. Rapamycin, an immunosuppressant agent, actively inhibits mTOR activity, curtailing cancer cell growth. Consequently, different compounds derived from sirolimus have been developed as cancer therapies and are undergoing investigation in clinical studies. While specific studies on mTOR’s role in pancreatic cancer were not uncovered in the search results, the roles of mTOR in other cancers suggest its potential as a target for malignant neoplasms of the pancreas. However, more focused research is essential to fully comprehend the mechanisms and potential therapeutic applications of mTOR in pancreatic cancer. For more detailed information, it is advisable to consult with a healthcare professional..[62]

The transcription factor and signal transducer STAT3stands out as a hopeful focus for pancreatic cancer.There is evidence supporting the notion that selectively addressing STAT3 with a small molecule inhibitor holds potential as a treatment strategy for pancreatic cancer. STAT3 assumes a significant significant involvement in influencing the pancreatic cancer tumor microenvironment (TME). Notably, research indicates that STAT3 in tumor fibroblasts contributes to the establishment creating an immune-suppressed environment in pancreatic cancer^2^. Cancer-associated fibroblasts (CAFs), among the most prevalent cell types in the pancreatic cancer stroma, exhibit context-dependent regulation of tumor progression within the TME. Therefore, comprehending tumor-promoting pathways, including STAT3, in CAFs is crucial for the advancement of more effective stromal-targeting therapies. Additionally, a review examining the interplay between STAT3 and pancreatic cancer suggests that leveraging STAT3 inhibitors in the setting of pancreatic cancer might open avenues for innovative chemotherapeutic modalities..[63] [64] [65]

Cyclin D1 (CCND1), Cyclin D1, a protein generated by the CCND1 gene, has a crucial function in regulating the cell cycle. regulation and is frequently elevated in different types of cancers, including pancreatic cancer. In the setting of pancreatic cancer, CCND1 can stimulate cell proliferation by facilitating the transition from the G1 to the S phase of the cell cycle. The heightened expression of CCND1 can result in uncontrolled cell division, a characteristic feature of cancer. Furthermore, CCND1 has been linked to chemotherapy resistance in pancreatic cancer, as its overexpression enhances the viability of cancer cells, reducing their susceptibility to the cytotoxic effects of chemotherapy. Consequently, targeting CCND1 presents a potential avenue to impede the growth of pancreatic cancer cells and overcome chemotherapy resistance. However, in-depth research is necessary to fully comprehend the mechanisms and explore the therapeutic applications of CCND1 in pancreatic cancer. For more detailed information, it is advisable to consult with a healthcare professional.[4]

Chymotrypsin-like elastase family member 3B (CELA3B) functions as an enzyme in the pancreas associated with digestive functions. Its expression is exclusive to the pancreas, and studies have explored its potential diagnostic value in distinguishing distinguishing pancreatic from non-pancreatic cancers and within differentiating acinar cell carcinoma from adenocarcinoma of the ducts. In healthy tissues, there was immunostaining for CELA3B observed solely in acinar cells and a subset of ductal cells in the pancreas. Among tumors, 75% of acinar cell carcinoma cases (12 out of 16) showed CELA3B immunostaining, with 37.5% exhibiting strong staining. This indicates that CELA3B could act as a possible focus for pancreatic neoplasms, particularly in acinar cell carcinoma. However, more specific research is required to comprehensively understand the mechanisms and explore potential therapeutic applications of CELA3B in pancreatic cancer. For detailed information, it is advisable to consult with a healthcare professional.[66]

The protein The Epidermal Growth Factor Receptor (EGFR), which typically facilitates cell growth, has been recognized as a promising focus in the treatment of pancreatic cancer. In the context of pancreatic cancer treatment, inhibitors like Erlotinib (Tarceva) can be combined with the chemotherapy drug gemcitabine, particularly for individuals with pancreatic cancer in an advanced stage. The efficacy of this blend may vary among patients. Notably, research by Xiaoting demonstrated a positive response to furmonertinib in a patient with EGFR-sensitive mutation and KRAS wild-type advanced pancreatic cancer. Despite disease progression after three initial treatment regimens, furmonertinib administration led to tumor shrinkage and a survival without progression of 4.7 months. Additionally, a study highlighted the development of an EGFR-targeted and gemcitabine-incorporated chemo gene for a comprehensive approach to combinatorial pancreatic cancer treatment. This strategy aims to integrate diverse therapeutic targets, potentially enhancing the responsiveness of pancreatic cancer to chemotherapy and working synergistically improving effectiveness against tumors.[67] [68]

Deoxycytidine kinase (DCK) serves as a crucial enzyme involved in the rescue pathway of nucleoside metabolism, playing a vital function in the activation of various prodrugs of nucleoside analogs utilized in cancer chemotherapy. In the context of pancreatic cancer, DCK emerges as a potential therapeutic target owing to its involvement in drug activation. Specifically, gemcitabine, a frequently employed chemotherapeutic agent for pancreatic cancer, relies on DCK for activation. Consequently, the levels and function of DCK within cancer cells may impact the efficacy of gemcitabine treatment. Furthermore, DCK has been linked to the emergence of resistance to treatments based on nucleoside analogs chemotherapy. Overcoming this resistance poses a significant challenge in pancreatic cancer treatment, and targeting DCK holds potential to address this issue.[69]

Prostate Stem Cell Antigen (PSCA) is a protein typically associated with an unfavorable prognosis in pancreatic cancer. However, recent studies indicate that PSCA might exhibit tumor-suppressive properties when located outside the cells. Research by Kexin and colleagues revealed that extracellular PSCA in Pancreatic Ductal Adenocarcinoma (PDAC) demonstrated tumor-suppressing potential, weakening cancer cells and diminishing their ability to spread. Additionally, PSCA was found to enhance the effectiveness of other anti-cancer medications. Notably, the incorporation of PSCA into pancreatic ductal adenocarcinoma (PDAC) cell lines resulted in reduced Mesothelin (MSLN) levels, suggesting that extracellular PSCA could exert anti-tumor actions, possibly through interactions with MSLN.[70]

Chymotrypsin-like elastase family member 1 (CELA1) is an enzyme in the pancreas responsible for digestive processes. CELA1 expression is restricted to the pancreas, and its possible diagnostic value has been explored for distinguishing pancreatic from extra-pancreatic cancers, as well as differentiating distinguishing acinar cell carcinoma from ductal adenocarcinoma. In healthy tissues, CELA1 there was immunostaining observed solely in acinar cells and a fraction of ductal cells in the pancreas. Among tumors, CELA1 staining for the immune system was detected in 12 out of 16 (75%) acinar cell carcinomas of the pancreas, including 6 cases with intense staining (37.5%). Consequently, CELA1 could potentially serve as a target for malignant neoplasms of the pancreas, especially in the context of acinar cell carcinoma. However, more specific additional investigation is needed to comprehensively comprehend the mechanisms and potential therapeutic applications of CELA1 in pancreatic cancer. It is advisable to consult with a healthcare professional for more detailed information.[3]

CDKN1A, also recognized as p21, has been identified as a possible focus for cellular immunotherapy in cancer treatment. Its pivotal role involves enhancing the capacity of macrophages derived from monocytes (MDMs) to engulf cancer cells. This process is achieved by transcriptionally repressing SIRPα (Signal-Regulatory Protein α), a gene responsible for encoding a phagocytic inhibitor. Consequently, these MDMs acquire an inflammatory pattern that reaches neighboring MDMs in a manner dependent on Interferon γ (IFNγ). It is essential to note that the referenced research studies are specific to leukemia and not directly related to pancreatic cancer. The mechanisms involved in pancreatic cancer may differ, and additional further investigation is required to validate the function of CDKN1A in this context particular context. [71]

Vascular endothelial growth factor A (VEGFA) and its receptor VEGFR-2 play a vital function in regulating tumor-induced angiogenesis. Both are present not only in endothelial cells but also in pancreatic cancer cells. VEGFA significantly enhances the movement of cancerous pancreatic cells, a process that can be suppressed by VEGFR-2 siRNA. The medium conditioned by pancreatic cancer cells notably boosts the motility of these cells. Inhibitors targeting VEGF/VEGFR, such as bevacizumab and sunitinib, effectively reduce The movement patterns of pancreatic cancer cells. VEGFA increases VEGFR-2 phosphorylation levels in pancreatic cancer cells. Bevacizumab and sunitinib decrease VEGFR-2 phosphorylation, along with reduced The expression of phosphorylated ERK (p-ERK) and phosphorylated Akt (p-Akt). Additionally, VEGFA lowers the expression of zonula occludens (ZO-1) or ZO-2 in pancreatic cancer cells. Hence, the signaling pathway involving VEGFA/VEGFR-2 plays a vital Contribution to the facilitation of invasion and migration in pancreatic cancer cell activity. This positions VEGFA as a potential target for treating malignant neoplasms of the pancreas. However, the current use of VEGFR inhibitors faces limitations, including restricted clinical efficacy and potential toxicity. Consequently, there is a need for the development of new strategies to enhance clinical results and reduce adverse effects to a minimum. associated with VEGFR inhibitors.[72] [73]

CD274, also known as te pivotal role in the immune system is performed by programmed cell death protein ligand 1 (PD-L1). checkpoint regulation and has demonstrated significant success in inducing tumor remissions across various human cancers. The expression of CD274 is primarily attributed to transcription factor (TF) activity, with the remaining unexplained instances largely associated with mutations or low microRNA abundance. Key regulators like IRF1, STAT1, NFKB, and BRD4 account for 90–98% of CD274 mRNA levels in patients. Within pancreatic ductal adenocarcinoma (PDAC), there is a notable frequency of CD274 loss, suggesting its possible as a focal point for addressing pancreatic malignancies. However, a substantial portion of patients shows resistance to treatment, underscoring the need for further research to comprehensively understand CD274 mechanisms and develop more effective therapeutic approaches. In addition to genetics-guided treatment, immunotherapies such as chimeric antigen receptor T cells (CAR-T), antibody-drug conjugates, and immune checkpoint inhibitors hold promise for precise tumor targeting. Despite these advancements, the clinical utility of immunotherapies in PDAC treatment remains limited. Therefore, while CD274 stands out as a hopeful focal point for treating pancreatic cancer, ongoing research aims to enhance the efficacy of CD274-targeting therapies and minimize potential toxic effects.[74]

CD24, a heavily glycosylated mucin-like compound, has undergone thorough exploration examined in the capacity of a marker for cancer stem cells across various solid cancers, including pancreatic cancer. Functioning as a “don’t eat me” signal, CD24 binds inhibiting macrophage phagocytosis by binding to Siglec-10. It serves as a marker for cancer cell stemness, influencing multiple signaling pathways associated with growth, infiltration, and spreading to other parts of the body. The existence of CD24 has demonstrated a strong association with unfavorable clinical outcomes in various tumor types, making it a robust candidate for T cell-engaging bispecific antibodies (BsAbs). Research suggests that a CD24-targeted antibody could effectively kill cancer cells and disrupt the recurrence cycle from cancer stem cells. While limited efficacy data exists, two clinical trials have confirmed the clinical safety and tolerability of monoclonal antibodies targeting CD24. Preclinical evaluations have also explored other modalities such as antibody-drug conjugates and chimeric antigen receptor (CAR) T cell therapy. In conclusion, CD24 emerges as a promising target for pancreatic cancer treatment. Nevertheless, ongoing research aims to enhance the efficacy of CD24-targeting therapies and minimize potential toxic effects.[75] [76]

Doublecortin-like kinase 1 (DCLK1) represents a protein recognized as a marker for tumor stem cells in gastrointestinal tract, pancreatic, and human cells associated with colorectal cancer. DCLK1 is linked to elevated. Expression of the KRAS gene through the pathway involving PI3K/AKT/mTOR. Consequently, targeting DCLK1 holds potential to reduce KRAS expression and regulate invasive behavior in pancreatic ductal adenocarcinomas (PDAC). Noteworthy variations in DCLK1 differences in expression between tissues of solid tumors and their adjacent normal tissues have been documented. In instances with low CD8+ T-cell infiltration, distinctive immunoreactive DCLK1 was noted. This suggests that DCLK1 could serve as a potentially favorable target for addressing pancreatic cancer.[77] [78]

CTNNB1, also referred to as beta-catenin, is a pivotal protein participating in regulating cell adhesion and transcriptional activity of genes. Mutations in the CTNNB1 gene have been detected in diverse cancer forms., including pancreatic cancer. Notably, studies focusing on solid-pseudopapillary neoplasms (SPNs), a rare pancreatic tumor type, have revealed frequent mutations occurring in exon 3 of the CTNNB1 gene. These mutations were detected through next-generation sequencing (NGS) on tissue samples obtained from SPNs. Additionally, a separate study detailed two cases of “pure” hepatoid tumors in the pancreas displaying somatic mutations in the CTNNB1 gene. These tumors exhibited common features and demonstrated indolent biological behavior. The findings suggested indicating that these tumors constitute a separate entity compared to pancreatic ductal adenocarcinoma (PDAC) and warrants recognition as a novel form of solid pseudopapillary neoplasms. Consequently, CTNNB1 emerges as a potential target for treating pancreatic cancer. Nonetheless, further research is essential to enhance the effectiveness of therapies targeting CTNNB1 and to minimize potential adverse effects.[79] [80]

MUC16, also referred to as CA125, stands out as a protein biomarker exhibiting significant overexpression in pancreatic cancer. It has been recognized as a promising potential choice for fluorescence-guided surgery (FGS) within the realm of pancreatic cancer. Recent investigations uncover MUC16 overexpression in 60-80% of pancreatic cancer cases. A near-infrared fluorescence antibody specific to MUC16 has been created for use in surgical imaging. This development holds potential for precise delineation of tumor tissue during surgery, facilitating complete resection and the identification of locally metastatic disease. Such precision can spare patients from unnecessary major surgeries, allowing them to proceed directly to alternative treatments. Additionally, a fluorescent antibody probe targeting MUC16, named AR9.6-IRDye800, has been designed for the resection of pancreatic cancer guided by imaging. The probe has shown effectiveness in binding to human pancreatic cancer cell lines in both laboratory settings and living organisms.[81] [82] [83]

Ribonucleotide reductase catalytic subunit M1 (RRM1) has been identified in datasets associated with gemcitabine. It has been observed that the expression of RRM1 is notably elevated in pancreatic cancer tissue as well as in cells resistant to gemcitabine in both laboratory and living organism settings settings. Knocking down Reversal of gemcitabine by RRM1 resistance, hindered migration and invasion. Additionally, individuals exhibiting elevated RRM1 levels levels experienced a reduced lifespan. A constructed nomogram based on RRM1 effectively predicted prognosis and underwent further validation. Consequently, RRM1 emerges as a possible indicator for predicting outcomes and a focal point for resistance to gemcitabine in pancreatic cancer. In a study by Kato et al, it was revealed RRM1 expression showed an elevation 24 hours post gemcitabine exposure, and this increase could be inhibited by blocking the activity of histone acetyltransferase. Activation of cytoplasmic. The reaction of RRM1 to gemcitabine exposure was primarily activated within the cytoplasm, and this activation was associated with the viability of cancer cells. Conversely, cancer cells without RRM1 activation within the cytoplasm exhibited significant DNA damage. Inhibition of RRM1 using specific siRNA or hydroxyurea augmented the toxic impact of gemcitabine on pancreatic cancer cells. In conclusion, RRM1 is intricately involved in biological processes associated with drug resistance following exposure to gemcitabine, positioning it as a possible focus for the treatment of pancreatic cancer. Nonetheless, further research is imperative to improve the effectiveness of treatments directed at RRM1 and reduce potential adverse effects. [84] [85]

MicroRNA-21 (miR-21) stands out as one of the highly expressed and widely prevalent researched microRNAs, exerting crucial regulatory functions in both healthy and diseased cells. Its involvement has been identified in various pathologies, spanning neoplastic and non-neoplastic conditions, acting through interactions with numerous gene targets. Among the primary The molecules targeted by miR-21 include Phosphatase and Tensin Homolog (PTEN), Tropomyosin 1 (TPM1), and Programmed Cell Death 4 (PDCD4). In the realm of digestive system cancers, miR-21 has undergone thorough investigation regarding its biological functions and therapeutic potential. Its roles as a biomarker for diagnosis and prognosis, along with its therapeutic utilizations, have undergone thorough exploration explored. However, it is crucial to acknowledge that miR-21 has been proposed as a biomarker for prediction or prognosis in a minimum of 29 diseases, lacking specificity to any particular ailment. Consequently, while miR-21 holds promise as a target for pancreatic cancer treatment, further research is imperative to enhance the effectiveness of therapies targeting miR-21 and to mitigate potential toxic effects.[86] [87]

Hypoxia-inducible factor-1 alpha (HIF-1α) plays a vital role as a regulator in cellular responses to changes in oxygen levels, facilitating the adjustment of cancer cells to hypoxic conditions within the oxygen-deficient tumor microenvironment. The induction of autophagy by HIF-1α has been identified as a contributor to transition from epithelial to mesenchymal states (EMT) and the enhanced movement of stem cells in pancreatic cancer, ultimately elevating the aggressive nature of pancreatic cancer. Hypoxia-inducible factor-1 alpha (HIF-1α)orchestrates malignant traits in pancreatic cancer through diverse pathways, suggesting that aiming at HIF-1α and the correlated signaling routes holds potential for therapeutic efficacy for pancreatic cancer. However, directly blocking HIF-1α is challenging due to its role as a transcription factor primarily located in the nucleus. Research by Tao et al has demonstrated that the KRAS/MEK/ERK signaling pathway exacerbates hypoxia-driven stabilization of HIF-1α and HIF-2α proteins in pancreatic ductal adenocarcinoma (PDAC) cells with activated KRAS, leading to the upregulation of subsequent effectors like CA9 and monocarboxylate transporter (MCT) 4. This process disrupts pH regulation and shifts metabolic patterns towards glycolytic phenotypes. In conclusion, HIF-1α is implicated in processes related to drug resistance under hypoxic conditions and represents a possible focal point for pancreatic cancer treatment. However, further research is essential to enhance the effectiveness of therapies targeting HIF-1α and to minimize potential toxic effects.[88], [89]

### 1.2. Malignant neoplasm of pancreas treatments

#### Targeted Therapy

Secretin, a hormone vital for maintaining water balance throughout the body, plays a regulatory role in the duodenal environment by controlling fluids released in the stomach, pancreas, and liver. Produced in the S cells of the duodenum, secretin is a biotech drug and a protein-based therapy. Its use involves stimulating pancreatic or gastric secretions for diagnosing issues such as dysfunction in the exocrine pancreas, gastrinoma, and irregularities in the bile and pancreatic ducts. Secretin helps evaluate the functionality of the pancreas, detect tumors in the pancreas or bowel, and its primary function is to prompt the pancreas to release pancreatic fluid for maintaining pH balance in the small intestine. Additionally, secretin contributes to body fluid homeostasis and bile production. When stomach hydrochloric acid enters the duodenum, secretin is released into the bloodstream, activating pancreatic duct cells to secrete water and bicarbonate. This process effectively dilutes and neutralizes the potentially damaging effects of stomach acid on the intestinal lining.[90]

Glucagon, a hormone produced by the pancreas’ alpha cells, is employed for therapeutic purposes severe hypoglycemia and as a tool for diagnosis in radiological exams to temporarily halt gastrointestinal tract movement. As a biotech drug and a peptide hormone, meaning it is a protein-based therapy, glucagon is utilized to elevate blood sugar levels by prompting the liver to convert stored glycogen into glucose. Additionally, it is administered to suspend stomach movement during radiological examinations for diagnosing specific stomach or intestinal disorders. The binding of glucagon to the glucagon receptor activates Gsα and Gq, leading to increased intracellular cyclic AMP and protein kinase A activation. The stimulation of Gq triggers phospholipase C, elevating inositol 1,4,5-triphosphate production and releasing calcium within cells. This process involves protein kinase A phosphorylating glycogen phosphorylase kinase, subsequently phosphorylating glycogen phosphorylase, and leading to the breakdown of glycogen.[91], [92], [93]

Azaserine, a serine derivative diazo compound found in nature, possesses both properties that act against both cancerous cells and bacteria. Classified as a small molecule, it has been utilized to impede the growth of certain tumors, showcasing anticancer activity. Despite its potential, the clinical effectiveness of Azaserine has shown variability. Mechanistically, Azaserine competitively inhibits glutamine amidotransferase, a pivotal enzyme in glutamine metabolism. This inhibition impacts It inhibits the rate-controlling stage of the metabolic hexosamine pathway and permanently blocks γ-glutamyl. transferase by directly influencing the pocket where the substrate binds. Notably, Azaserine has also demonstrated the ability to safeguard against hyperglycemic endothelial damage by elevating levels of manganese-superoxide dismutase in the bloodstream, resulting in a direct decrease in the level of reactive oxygen species.[94], [95]

Therapeutic insulin, primarily employed in managing diabetes characterized by elevated blood sugar levels, is not commonly utilized directly for addressing pancreatic cancer. Ongoing research explores the involvement of insulin and insulin-like growth factors in the onset and progression of pancreatic cancer. As a biotech drug, therapeutic insulin is a protein-based hormone used to regulate glucose levels and facilitate glucose storage in the liver. While not a direct treatment for pancreatic cancer, it is noteworthy that individuals with diabetes, especially type 2 diabetes, face a heightened likelihood of developing pancreatic cancer. Insulin functions by enhancing glucose uptake and metabolism in cells, reducing blood sugar levels, and supporting energy production in cells. Additionally, it aids in storing excess glucose in the liver for future use, contributing to maintaining blood sugar levels within a healthy range. [96], [97], [98], [99], [100]

Pancreatic enzymes constitute a cluster of digestive enzymes crucial for breaking down food in the digestive system. While not commonly employed as a direct treatment for pancreatic cancer, they find utility in managing symptoms associated with pancreatic insufficiency resulting from pancreatic diseases or treatments. As biotech drugs, these protein-based therapies, specifically enzymes, are components of therapy involving the replacement of pancreatic enzymes (PERT). PERT is utilized to address pancreatic insufficiency, a condition where the pancreas fails to produce sufficient enzymes for proper digestion, often seen in pancreatic cancer. By enhancing digestion and absorption of nutrients, PERT mitigates symptoms such as weight loss, indigestion, and post-meal cramping. The pancreatic enzymes encompass amylase, which breaks down carbohydrates into sugars, protease, responsible for breaking down proteins into amino acids, and lipase, which facilitates the breakdown of fats into fatty acids and glycerol. Subsequently, these smaller molecules are absorbed through intestinal cells into the bloodstream.[101]

Everolimus is a medication used for the management of diverse forms of cancer, including advanced pancreatic neuroendocrine tumors. Everolimus is a small molecule drug. Everolimus is employed as a targeted therapy for advanced pancreatic neuroendocrine tumors. It has demonstrated anti-tumor efficacy in individuals with these tumors in phase 2 trials. Everolimus demonstrated a median progression-free survival of 11.0 months, contrasting with 4.6 months observed with a placebo. Everolimus works by inhibiting a protein called target of rapamycin in mammals (mTOR). mTOR is a type of kinase protein governing cell growth and metabolism. By inhibiting mTOR, Everolimus can block the progression of cells from the G1 phase to the S phase, thereby reducing the growth of tumors.[102], [103], [104], [105], [106]

Triptolide, characterized as a small molecule, has demonstrated promise as an innovative therapeutic option for pancreatic cancer. Research has explored its anti-tumor effectiveness in various pancreatic cancer cell lines and cells obtained directly from patients those with cells obtained directly from patients. The mechanism of action involves targeting several proteins, such as polycystin-2, ADAM10, DCTPP1, TAB1, and XPB. Triptolide exhibits pleiotropic effects, causing a reduction in The expression of heat shock protein 70 (HSP70), influencing release of calcium, inducing depolarization of the lysosomal membrane, suppressing the activity of NFκB, suppressing iNOS and Cox-2 expression, acting as a inhibitor of transcription, and serving as a factor that inhibits angiogenesis. A new pro-drug of Triptolide, (E)-19-[(1’-benzoyloxy-1’-phenyl)-methylidene]-Triptolide (CK21), has been synthesized and prepared as an emulsion for testing both in laboratory settings and living organisms. CK21 demonstrates demonstrates strong anti-proliferative effects on human pancreatic cancer cell lines and pancreatic tumor organoids derived from patients in laboratory settings, while exhibiting minimal toxicity in living organisms. Its mode of action involves the inhibition of the NF-κB pathway, ultimately resulting in mitochondrial-mediated apoptosis of tumor cells.[107], [107], [108], [108], [109], [110]

#### Chemotherapy

Gemcitabine, a chemotherapy agent utilized for the management of different types of cancers, including pancreatic malignancies, is classified as a small molecule drug and is a member of the nucleoside analog category. Its mechanism of action involves hindering the production of fresh DNA, leading to cell death. Upon entry into cancerous cells, gemcitabine undergoes phosphorylation Deoxycytidine kinase converts it into gemcitabine monophosphate, which is then transformed into active compounds known as gemcitabine diphosphate (dFdCDP) and gemcitabine triphosphate (dFdCTP). Typically administered as a A 1,000 mg/m^2^ dose administered through a 30-minute infusion once a week for three weeks within a four-week cycle. recent studies indicate that combining gemcitabine with other therapies can enhance its efficacy. Long-term gemcitabine treatment has been shown to reshape the microenvironment of pancreatic tumors and sensitize applying combination immunotherapy to carcinoma. Additionally, a novel gemcitabine analog, 4-(N)-stearoyl-gemcitabine (4NSG), has demonstrated improved systemic stability and increased antitumor efficacy in xenograft models derived from patients with pancreatic cancer. [3],[111],[112],[113], [114],[115],[116],[117]

FOLFIRINOX, a chemotherapy regimen employed for the treatment of advanced pancreatic cancer, constitutes a blend of four small molecule drugs. These include Folinic acid (Leucovorin), a vitamin B derivative amplifying the effects of 5-fluorouracil (5-FU); Fluorouracil (5-FU), a pyrimidine analog and antimetabolite that becomes part of the DNA structure, halting Irinotecan inhibits the uncoiling of DNA by acting on topoisomerase, interfering with DNA synthesis. and duplication; and Oxaliplatin, a platinum-based antineoplastic agent inhibiting DNA repair and/or synthesis. The collaborative action of these drugs targets cancer cells through diverse mechanisms. 5-FU disrupts DNA synthesis, irinotecan impedes DNA uncoiling and duplication, and oxaliplatin hinders DNA repair and/or synthesis. Administered every 14 days for a maximum of 12 cycles, FOLFIRINOX has demonstrated the ability to extend survival in advanced pancreatic cancer patients, with a no table 4-month improvement compared to the standard gemcitabine treatment. Despite its efficacy, FOLFIRINOX is associated with substantial toxicity and serious side effects, making it suitable only for patients with a favorable performance status.[118]

Both FOLFIRINOX and Modified FOLFIRINOX (mFOLFIRINOX) are chemotherapy protocols employed in pancreatic cancer treatment, featuring the same drug components: Folinic acid (Leucovorin), Fluorouracil (5-FU), Irinotecan, and Oxaliplatin. The primary distinction lies in dosing and administration. FOLFIRINOX, the original regimen, involves a bolus administration of 5-FU, contributing to significant toxicity and potential limitations in patients with poor functional status. Conversely, Modified FOLFIRINOX (mFOLFIRINOX) excludes the bolus administration of 5-FU, aiming to mitigate toxicity. Studies have indicated comparable efficacy against metastatic pancreatic adenocarcinoma (MPA) between mFOLFIRINOX and standard FOLFIRINOX. However, mFOLFIRINOX is linked to a lower rate of dose decrease but a marginal elevated frequency of severe adverse effects.[119]

Fluorouracil, also referred to as 5-FU, is a chemotherapy substance utilized for managing different types of cancers, which encompasses malignant neoplasm of the pancreas. Classified as a small molecule drug, it belongs to the antimetabolite and pyrimidine analog drug categories. The mode of operation entails inhibiting the synthesis of DNA and proteins, essential for the proliferation of cancer cells. Functioning as a synthetic analogue of pyrimidine, Fluorouracil undergoes conversion to multiple active metabolites in the body. These metabolites integrate into DNA and RNA, disrupting their structures and impeding protein synthesis. This disruption hinders the growth and division of cancer cells and other rapidly dividing cells, ultimately leading to their demise. Administration of Fluorouracil is typically intravenous, incorporated into cycles of treatment with intermittent rest periods to allow the body to recover. The frequency of administration varies based on cancer type, occurring weekly or in cycles every 2, 3, or 4 weeks. In some cases, continuous treatment is administered through a portable pump. Recent studies indicate that Fluorouracil’s efficacy can be augmented when used in combination with other therapies. For example, individuals with recurring pancreatic cancer undergoing systemic therapy, including the standard combination of A median was shown in a regimen involving 5-fluorouracil, irinotecan, and oxaliplatin. survival rate of 14 months.[120],[121],[122],[123]

The GNP regimen, which denotes The combination of gemcitabine and nab-paclitaxel constitutes a chemotherapy regimen approach utilized for treating advanced pancreatic cancer. It involves two small molecule drugs: Gemcitabine, a a pyrimidine analog and antimetabolite compound that becomes part of the DNA structure, impeding the creation of DNA; nab-Paclitaxel, a unique version of the antineoplastic agent paclitaxel bound to albumin, inhibiting cell division. The collaborative action of these drugs results in the synergistic elimination of cancer cells through distinct mechanisms. Gemcitabine halts DNA synthesis, and nab-Paclitaxel impedes cell division. Administered every 14 days for a maximum of 12 cycles^1^, recent studies indicate that GNP can prolong the viability of individuals having progressed pancreatic cancer. Those subjected to GNP survived approximately 10.3 months, compared to 11.8 months for FOLFIRINOX recipients. However, GNP is a potentially highly toxic drug combination with significant side effects, and only individuals with a positive functional status eligible for this regimen.[124],[125],[126],[127],[128],[129]

Irinotecan is a small molecule medication employed in treating various cancers, including pancreatic cancer. As a topoisomerase I inhibitor, Irinotecan functions by obstructing the activity of the topoisomerase I enzyme crucial for cell division and growth. Its mechanism involves preventing the reconnection binding to the DNA strand the topoisomerase I-DNA complex, leading to DNA damage and subsequent cell death. The primary use of Irinotecan is in colorectal cancer therapy, particularly when used in conjunction with other chemotherapy agents, such as the FOLFIRI regimen comprising 5-fluorouracil, leucovorin, and irinotecan. Additionally, it may be employed in combination with fluorouracil and folinic acid for pancreatic cancer following the initial therapy. failure. Recent studies highlight the intricate metabolism of irinotecan, involving its breakdown through the action of carboxylesterases into the active form known as SN-38, significantly more potent than irinotecan itself. Genetic variations within the DNA of these enzymes and transporters have been identified as predictors of drug-related toxicity and treatment efficacy, as demonstrated in reviews of past and future trials and comprehensive analyses.[3], [120], [130], [131], [132]

Oxaliplatin is a small molecule medication utilized in treating various cancers, including pancreatic cancer. As a platinum drug and alkylating agent, Oxaliplatin functions by impeding DNA synthesis in cells, halting or slowing the growth of cancer cells and other rapidly dividing cells, ultimately leading to their demise. In physiological solutions, it undergoes nonenzymatic conversion to active derivatives by displacing the labile oxalate ligand, generating transient reactive species like monoaquo and diaquo DACH platinum that covalently bind with macromolecules. Oxaliplatin is primarily employed in colorectal cancer therapy, particularly when used in conjunction with additional approaches chemotherapy substances, such as the FOLFIRI regimen comprising 5-fluorouracil, leucovorin, and irinotecan. It may also be used in conjunction in combination with fluorouracil and folinic acid for pancreatic cancer following the failure of initial treatment. Recent studies reveal the intricate metabolism of oxaliplatin, involving its hydrolysis by carboxylesterases into the active metabolite SN-38, significantly more potent than oxaliplatin itself. Genetic variations within the genetic material of these enzymes and transporters have been identified as predictors of drug-related toxicity and treatment efficacy, demonstrated in both examinations of past and future trials, as well as comprehensive analyses.[3], [133], [133], [134], [135], [136]

Paclitaxel, sold under the trade name Taxol and others, is a chemotherapy agent employed for different cancer types, including pancreatic cancer^6^. Classified as a small molecule medication, it falls under the drugs classified as taxanes. Paclitaxel disrupts the normal function of tubular structures in cell division, facilitating microtubule assembly, stabilizing existing microtubules, and hindering their disassembly^5^. This disruption occurs during the late G2 mitotic phase, hindering cell replication. Administered intravenously^6^, Paclitaxel is notably myelosuppressive. For individuals with limited If there has been extensive prior treatment, the presence of bone metastasis, or significant skeletal radiation leading to depleted bone marrow reserves, an initial dosage of 60mg/m2 may be considered, potentially increased to 80mg/m2 if well tolerated. Given in a 7-day cycle for 18 cycles, recent studies highlight Paclitaxel’s potential in combination with other therapies to enhance effectiveness. Noteworthy is the well-tolerated pairing TTFields with nab-paclitaxel and gemcitabine (GnP), as demonstrated in the PANOVA phase 2 study, showing promising efficacy in pancreatic adenocarcinoma that has spread or is locally advanced.[120], [129], [137], [137], [138], [139], [140], [141]

Erlotinib, marketed as Tarceva, is a pharmaceutical agent used in the management of various cancers, including pancreatic cancer^16^. Categorized as a tyrosine kinase inhibitor, erlotinib acts by blocking the kinase function of the epidermal growth factor receptor (EGFR), a protein crucial for the growth and survival of cells. EGFR is present on the outer layer of both regular and cancer cells. Excessive EGFR in some cancer cells leads to uncontrolled growth. Erlotinib’s blockade of EGFR can result in the shrinkage or temporary cessation of cancer growth. Its primary use is in treating non-small cell lung cancer (NSCLC) and pancreatic cancer. in particular for NSCLC, it is reserved for cases with specific mutations in their EGFR protein. In pancreatic cancer, erlotinib is combined with gemcitabine. Most trials included in studies utilized a daily oral administration of 150mg of erlotinib. Recent research indicates that erlotinib can be employed synergistically with other therapies to augment its effectiveness. For example, studies have explored the impact of neoadjuvant therapy (gemcitabine/gemcitabine + erlotinib/gemcitabine + oxaliplatin/FOLFIRINOX/other regimens) in enriching antitumor immune cells within the tumor microenvironment.[3], [109], [142], [143], [144], [145], [146], [147, p. 3]

Oxonic Acid, also recognized as Oteracil, is a small molecule drug employed alongside other chemotherapy agents for managing advanced gastric cancer^12^. It serves as an adjunct to antineoplastic therapy, playing a crucial role in Teysuno (a commercially available product containing Oxonic Acid). The primary function of Oxonic Acid in Teysuno is to diminish the functioning of 5-FU in the regular gastrointestinal lining, thereby mitigating gastrointestinal toxicity. This is achieved by inhibiting the enzyme orotate phosphoribosyl transferase (OPRT, which plays a crucial role in the synthesis of 5-FU. Oxonic Acid is prescribed as an adjunct to antineoplastic therapy^12^. When integrated into Teysuno, Oxonic Acid is specifically indicated for treating adults with advanced gastric (stomach) cancer in combination with cisplatin. Recent research indicates that Oxonic Acid’s effectiveness can be enhanced when used in conjunction with other therapies.[126]

Tegafur is a chemotherapy medication employed for the management of different types of cancers, encompassing pancreatic cancer. As a small molecule drug and a member of the pyrimidine analogues class, Tegafur acts as a prodrug for 5-fluorouracil (5-FU), an antineoplastic agent^3^. Upon conversion to 5-FU and subsequent bioactivation, the drug exhibits It demonstrates anti-cancer effects by hindering thymidylate synthase (TS) in the pyrimidine pathway. crucial for DNA synthesis^3^. This interference with DNA replication impedes the growth of cancer cells and other rapidly dividing cells, leading to their demise. Tegafur is frequently administered in conjunction with additional medications for pancreatic cancer treatment, such as Gimeracil and Oteracil or in conjunction with Fluorouracil as Tegafur-uracil^3^. Typically, Tegafur is paired with other drugs that either enhance 5-FU bioavailability by inhibiting its degrading enzyme or mitigate 5-FU toxicity while maintaining high concentrations at a lower tegafur dose^3^. A case report illustrated the effectiveness of transitioning to a combination of TS-1 chemotherapy and anlotinib targeted therapy in a patient diagnosed with locally advanced pancreatic cancer.[150], [151], [151], [152]

The GEM/CAP regimen, incorporating gemcitabine and capecitabine, is employed for the treatment of pancreatic cancer. Both gemcitabine and capecitabine are classified as small molecule drugs. This regimen serves as both an adjuvant and initial therapy for advanced pancreatic cancer. Typically, the treatment is administered in cycles spanning a few months, with each cycle lasting 4 weeks. Patients commonly undergo six cycles of treatment over a 6-month period. Gemcitabine and capecitabine, as chemotherapy medications, focus on swiftly proliferating cells, such as cancer cells. Gemcitabine is administered intravenously, while capecitabine is taken orally. Within cancer cells, capecitabine undergoes conversion to 5-fluorouracil (5-FU), the active form of the medication.[153], [154]

The GemOx protocol, comprising gemcitabine and oxaliplatin, is utilized for the treatment of different types of cancers, including pancreatic cancer. Both gemcitabine and oxaliplatin fall into the category of small molecule drugs^5^. This regimen is employed for managing advanced pancreatic cancer, typically administered in cycles spanning a few months. Patients undergo administering gemcitabine at a dosage of 1000 mg/m via infusion administering oxaliplatin at a dosage of 100 mg/m via infusion on the second day. The therapy is reiterated every 2 weeks for a total of six cycles. Gemcitabine and oxaliplatin, acting as chemotherapeutic agents, focus on swiftly proliferating cells, including cancer cells. Gemcitabine, an antimetabolite, disrupts DNA synthesis, while oxaliplatin, an alkylating agent based on platinum, triggers alterations in DNA damage.[155], [156], [157]

Cisplatin is a chemotherapy medication employed in the management of diverse types of cancer, including pancreatic cancer. This small molecule drug is administered through intravenous brief infusion in standard saline for the management of both solid tumors and blood-related cancers. Cisplatin is utilized in the treatment of sarcomas, certain carcinomas (such as conditions such as small cell lung cancer, head and neck squamous cell carcinoma, ovarian cancer, lymphomas, bladder cancer, cervical cancer, and germ cell tumors. Its extensive application has significantly increased increasing the recovery rate for testicular cancer from 10% to 85%. Operating as an alkylating agent^2^, Cisplatin, containing the metal platinum, functions by attaching to DNA and hindering its replication. This unique mechanism of action damages the DNA of dividing cells irreparably, halting or slowing the growth of cancer cells and other rapidly dividing cells, leading to their demise.[158], [159], [160], [160], [161]

#### Immunotherapy

Capecitabine, available under the brand name Xeloda and others, is a chemical compound utilized in the therapy of various cancers, which encompasses malignant neoplasm of the pancreas. Classified as an antimetabolite, Capecitabine undergoes metabolism in the body to produce 5-fluorouracil (5-FU). This active metabolite, 5-FU is an antimetabolite compound. that disrupts the creation of DNA, RNA, and proteins, thereby impeding or slowing the proliferation of cancerous cells and other rapidly dividing cells, ultimately leading to their demise. Capecitabine is utilized either alone or in combination with other cancer treatments for individuals diagnosed with stage III colon cancer to prevent recurrence after undergoing surgical interventions. It is also administered alongside other cancer treatments and radiation therapy, typically around the time of surgery, to treat metastatic rectal cancer^3^. within the framework of pancreatic cancer, Capecitabine is employed in conjunction with other chemotherapy drugs. The common treatment strategy involves the oral administration of 150mg of capecitabine daily, as observed in most included trials. Recent studies have indicated that Capecitabine can be effectively combined with other therapies to enhance its overall effectiveness. For example, neoadjuvant therapy, encompassing various regimens like gemcitabine, gemcitabine with erlotinib, gemcitabine alongside oxaliplatin, and FOLFIRINOX, and other combinations including radiotherapy, has demonstrated a positive impact enhancing immune cells with anti-tumor properties in the context of tumor microenvironment.[3], [109], [162]

#### Antiviral Medication

Tegafur-gimeracil-oteracil potassium, commonly known as S-1, is a blend of three small molecule drugs: Tegafur, an antineoplastic prodrug of fluorouracil (5-FU) belonging to the ‘anti-metabolites’ group; Gimeracil, an enzyme inhibitor that blocks the reversible breakdown of fluorouracil inhibiting the dihydropyrimidine dehydrogenase (DPD) enzyme; Oteracil, an enzyme inhibitor that hinders the orotate phosphoribosyl transferase (OPRT) enzyme involved in 5-FU production. Tegafur transforms into fluorouracil more substantially more pronounced in tumor cells compared to normal tissues. Fluorouracil, resembling pyrimidine, a component of cellular genetic material (DNA and RNA), takes the place of pyrimidine in the body, disrupting enzymes crucial for new DNA synthesis. Consequently, it impedes tumor cell growth and leads to their eventual demise^2^. Gimeracil and Oteracil complement Tegafur’s activity by enhancing its effectiveness. Tegafur-gimeracil-oteracil potassium is prescribed for It is employed in conjunction with cisplatin for advanced gastric cancer^2^. Additionally, it is used in the therapy of head and neck, colorectal, non-small-cell lung, breast, pancreatic, and biliary tract cancers. Recent studies indicate that Tegafur-gimeracil-oteracil potassium can be effectively combined with other therapies to augment its efficacy.[163], [164], [165], [166]

#### Diagnostic

CA19-9 is a protein typically present on certain cell surfaces and is recognized as a tumor marker, serving as a monitoring tool for specific cancers and their treatment progress. While most commonly associated with pancreatic cancer, CA19-9 is also linked to other cancers like colon, stomach, bile duct, ovarian, and bladder cancer. It functions as Cancer Antigen 19-9, also known as Sialylated Lewis A, a tetrasaccharide attached to O-glycans on cell surfaces, contributing to cell-to-cell recognition processes. Primarily utilized in managing pancreatic cancer, CA19-9 is not employed as a standalone diagnostic tool but rather to assess cancer response to therapy or potential recurrence post-treatment. Monitoring CA19-9 levels aids in evaluating tumor secretion, indicating a response to treatment with subsequent declines and potential rises upon disease recurrence. It proves valuable as a surrogate marker for relapse. While CA19-9 is employed in cancer management, its levels can indicate cancer growth or shrinkage, aiding in predicting cancer behavior, detecting post-treatment recurrence, and assisting in the diagnosis of certain cancers and diseases when combined with other tests. Notably, recent studies highlight the potential use of CA19-9-targeted antibodies derived from CA19-9/keyhole limpet hemocyanin (KLH) vaccine-immunized patients in safeguarding rodents from pancreatic advancement of cancer, suggesting a potential role in vaccine therapy for pancreatic cancer. [167],[168],[169],[170],[171],[172]

Somatostatin, a peptide hormone with regulatory functions in the endocrine system, influencing neurotransmission and cell proliferation^6^, is not itself a drug but has clinical applications in diagnosing acromegaly and gastrointestinal tract tumors. Analogues of somatostatin have been developed to enhance efficiency in managing acute conditions like esophageal varices. As a small molecule, somatostatin is a peptide hormone that occurs naturally consisting consisting of either 14 or 28 amino acid units. Attaching to five subtypes of somatostatin receptors (SSTRs), which are transmembrane receptors coupled with Gi proteins, thereby suppressing adenylyl cyclase, somatostatin exhibits anti-neoplastic effects on tumors through direct, indirect, or combined actions. Somatostatin analogues (SSAs) are drugs addressing symptoms of carcinoid syndrome, mitigating diarrhea and flushing while impeding tumor growth. Recent prospective clinical trials exploring high-dose SSA and a case report on octreotide-LAR treatment for metastatic non-functioning pancreatic tumors indicate reduced tumor growth and progression.[173], [174], [175], [176], [177], [178], [179]

Pentetreotide, a drug utilized for visualizing somatostatin receptor-positive neuroendocrine tumors, is a small molecule. Functioning as an inhibitor of somatostatin receptors, it comes provided in a package with Indium-111and serves used as a contrast medium for imaging somatostatin receptor-positive neuroendocrine tumors, including specific pancreatic tumors. While the complete mechanism of action is not thoroughly outlined in available sources, Pentetreotide is recognized for its ability to inhibit somatostatin receptors^1^. Somatostatin, a hormone inhibiting the release of various hormones like growth hormone, thyroid-stimulating hormone, and insulin, is affected by Pentetreotide through the inhibition of somatostatin receptors. This impact may influence the growth and behavior of certain cell types, including those present in neuroendocrine tumors.[180]

Amylase, an enzyme responsible for the hydrolysis of starch into sugars, is naturally present in the saliva of humans and some other mammals, initiating the digestive process. Although amylase is primarily not utilized as a drug for treating pancreatic malignancy, it serves a crucial role in the diagnostic assessment of pancreatic diseases by being measured in blood or pancreatic cyst fluid. As a biotech drug, specifically a protein-based therapy in the form of an enzyme, amylase is recognized for breaking down long-chain carbohydrates (starches) into smaller molecules. This enzymatic action facilitates the absorption of nutrients by the body. Elevated or reduced levels of amylase in diagnostic tests may indicate pancreatic issues, underlining its significance in assessing pancreatic health. Its mechanism involves the hydrolysis of alpha 1-4 connections in polysaccharides containing three or more glucose units linked together.[181], [182]

Somatostatin receptor agonists, a category of medications, have been employed in treating various tumors, including pancreatic neuroendocrine tumors. These agonists, characterized as small molecule drugs, interact with somatostatin receptors found on the cells of neuroendocrine tumors. Their successful application has led to a reduction in hormone secretion and an improvement in associated symptoms. It is important to note, however, that current evidence does not endorse the use of somatostatin receptor analogues for treating pancreatic neuroendocrine carcinoma. The mode of operation entails replicating the structure of neuropeptide somatostatin, targeting G protein-coupled receptors. Activation of these receptors prompts a decrease in hormone secretion within cells where the receptors are expressed. This process can impact neurotransmission and memory formation in the central nervous system. In both human and animal models, these agonists have demonstrated their effectiveness in inhibiting angiogenesis and reducing the proliferation of both healthy and cancerous cells.[177], [183], [184], [185], [186]

#### Other treatment strategy

Octreotide, categorized as a somatostatin analog, functions similarly to a natural hormone in the body, blocking the release of various hormones like growth hormone, insulin, and glucagon. This peptide drug, a synthetic analogue of somatostatin, is employed to address symptoms associated with specific tumors. As a small molecule drug and peptide hormone akin to somatostatin, octreotide binds to receptors on cells responsive to somatostatin, hindering their activity. This action diminishes the secretion of growth hormone released by the pituitary gland and insulin-like growth factor-1 (IGF-1) produced by the liver, alleviating metabolic and other symptoms of acromegaly. Octreotide is primarily used to treat symptoms linked to tumors, including carcinoid tumors and VIPomas, administered via injection and often integrated into a cyclical treatment plan with rest periods for the body to recover. The frequency of administration varies depending on the cancer type^8^, and in certain instances, continuous treatment may be facilitated through a portable pump. Recent studies highlight the affinity of Octreotide nanoparticles for in vivo induction of pancreas ductal adenocarcinoma using MIA Paca-2, indicating its potential utility in targeted therapy for pancreatic cancer.[121], [178], [187], [188]

Leucovorin, classified as a folic acid analog, shares similarities with the vitamin folic acid, crucial for new cell production and maintenance. When combined with specific chemotherapy drugs, Leucovorin enhances their efficacy in eradicating cancer cells or mitigating adverse side effects. Its mechanism involves safeguarding healthy cells from the impact of medications like methotrexate, enabling the entry of methotrexate to target and eliminate cancer cells. As a folic acid derivative, it can elevate folic acid levels, particularly after exposure to folic acid antagonists like methotrexate.Primarily used to alleviate the side effects of high doses or accidental overdose of medications impeding folic acid effects, including methotrexate, pyrimethamine, and trimethoprim, Leucovorin is administered through injection and is often integrated into multi-cycle treatment plans with intervals for body recovery. The frequency of administration varies based on cancer type, with some cases involving continuous treatment through a portable pump. Recent studies indicate that Leucovorin, when combined with other therapies like liposomal irinotecan and levoleucovorin for the treatment of pancreatic cancer, exhibited a reduction in tumor growth and progression.[3], [109], [152], [189], [190], [191]

Deoxycytidine, a deoxyribonucleoside integral to deoxyribonucleic acid, closely resembles the ribonucleoside cytidine, differing by the absence of one hydroxyl group at the C2’ position. This characteristic renders it a small molecule drug. When phosphorylated by deoxycytidine kinase, it transforms into deoxycytidine monophosphate (dCMP), a precursor to DNA. The ensuing conversion of dCMP to dUMP and dTMP highlights its role in DNA synthesis. Additionally, deoxycytidine serves as a precursor for 5-aza-2′-deoxycytidine, employed in treating myelodysplastic syndrome (MDS) by impeding the addition of methyl groups to the P15/INK4B gene. This intervention increases P15/INK4B protein expression, suppressing the progression of MDS to leukemia. Recent findings indicate that pancreatic stellate cells (PSCs) release deoxycytidine, contributing to gemcitabine resistance. Examination of metabolites in the media derived from distinct mouse PSCs revealed deoxycytidine’s role in inducing chemoresistance in pancreatic ductal adenocarcinoma (PDAC) cells by diminishing deoxycytidine kinase (dCK) capacity for gemcitabine phosphorylation^6^. However, human model data is lacking, and dCK expression didn’t show an association with gemcitabine’s clinical efficacy.[192], [193], [194], [195], [109], [196]

Diazoxide, a small molecule, stands out as an effective medical intervention for insulinoma, a specific type of pancreatic tumor. It is employed to address hypoglycemia associated with hyperinsulinism, often stemming from conditions like inoperable islet cell adenoma or carcinoma, as well as extrapancreatic malignancies. Diazoxide’s mechanism of action involves activating Potassium channels sensitive to ATP on the membrane of pancreatic beta-cells. This activation induces potassium efflux from beta-cells, resulting in reduced calcium influx and, subsequently, decreased insulin release. This mechanism proves valuable in treating hyperinsulinemic hypoglycemia due to its inhibitory effect on insulin release. However, it’s crucial to be aware of potential side effects when using Diazoxide. Reported adverse effects include fluid retention and electrolyte disturbances.

A study occurred more commonly observed in females than in males, and some patients exhibited unacceptable thrombocytopenia shortly after Diazoxide administration. While Diazoxide holds promise treating specific pancreatic tumors, consulting a healthcare professional is essential to grasp potential benefits and risks. Personalized advice determined by the status of the patient and overall health status can be provided by healthcare professionals. It’s crucial to emphasize that this information is derived from the latest available data and may evolve with new research findings.[197], [198], [199]

### 1.3. Objective

While numerous investigations have explored effective medications for Malignant neoplasm of pancreas, the factors delineated in Figure 1 have resulted in a dearth of comprehensive statistical analysis in this area. Conversely, artificial intelligence has garnered attention across diverse medical realms in recent times, ranging from protein folding and medical imaging to cohort studies and fundamental alterations in neural networks. [200], [201], [202], [203], [204]

**Figure 1:**
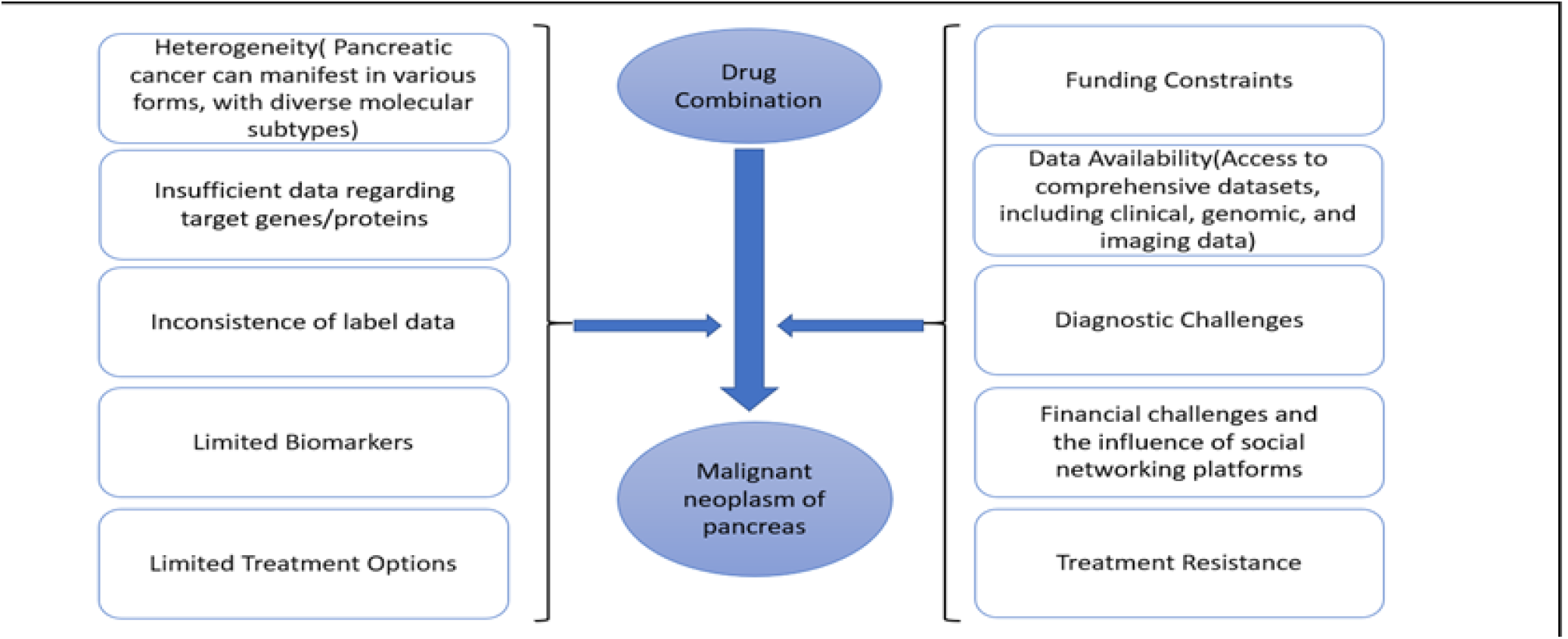
The effects of proposed drug combinations on the management of Malignant neoplasm of pancreas.

Recent publications introduce the RAIN protocol as a novel approach, involving the combination of drug associations to disease treatment, evaluating by p-value metric. The p-value indicating the association between the disease and target proteins/genes is near one. These papers employ various Artificial Intelligence algorithms, including Graph Neural Network for the suggestion of drug combinations. The usual practice involves conducting a network meta-analysis to evaluate comparative effectiveness.[205], [206], [207], [208], [209], [210], [211].

The RAIN protocol functions in the capacity of a comprehensive review and meta-analysis strategy, utilizing the STROBE method to tackle a specific medical question. [212], [213], [214]

## 2. METHODS

The RAIN protocol consists of three clear-cut phases. Initially, artificial intelligence is utilized to recommend the most effective combination of medications tailored for addressing and managing a particular condition. Following this, an exhaustive examination is carried out via Natural Language Processing, methodically scrutinizing recent publications and clinical trials to gauge the efficacy of different variations of the recommended combination. Figure 2 illustrates the breakdown of articles across each step of the STROBE checklist. Lastly, in the third phase, the efficacy of drugs and their linked human proteins/genes is assessed using Network meta-analysis. The subject also introduces the RAIN protocol, which encompasses Recommendation, Analysis,

**Figure 2:**
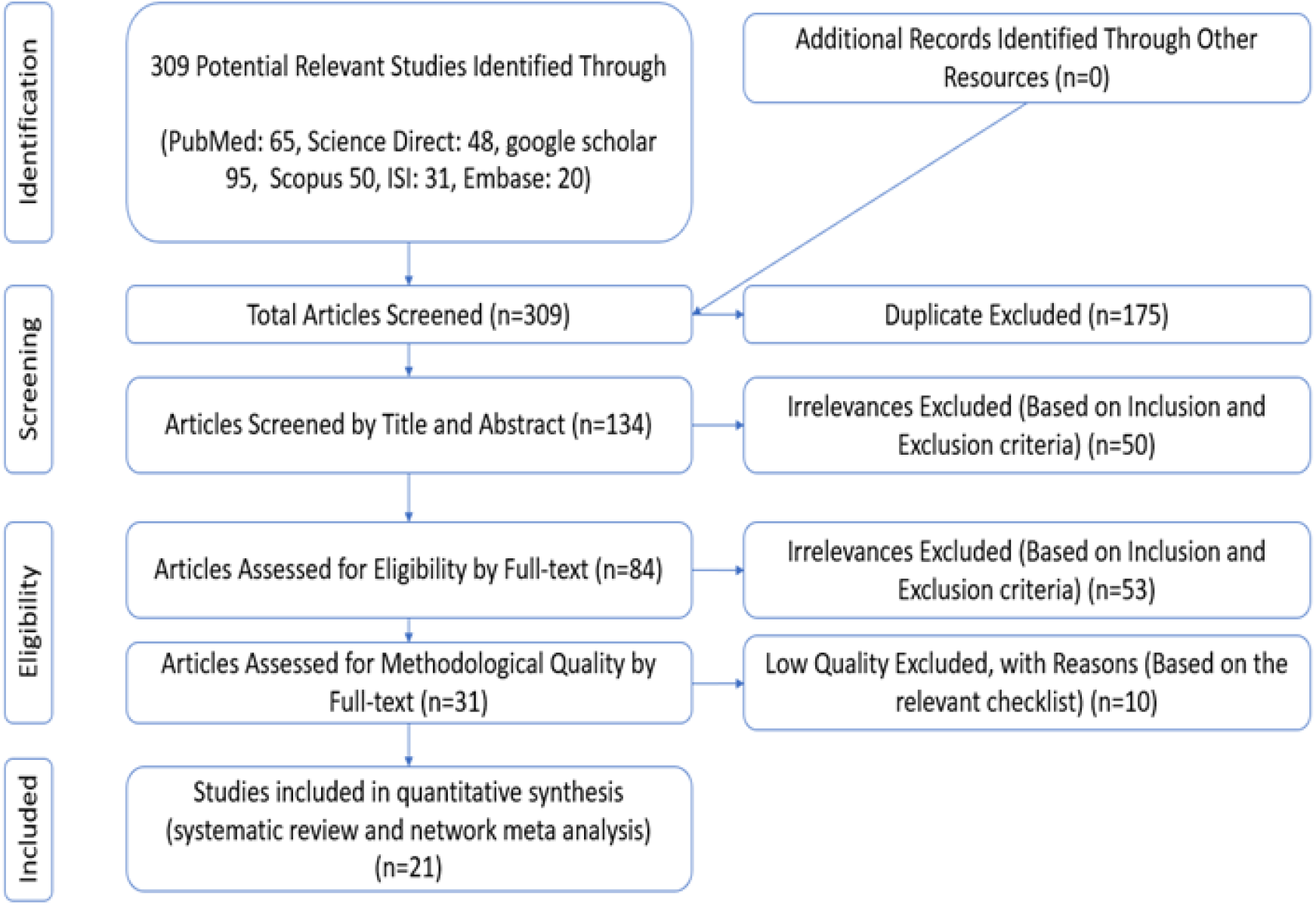
PRISMA (2020) flow diagram indicating the stages of sieving articles in this RAIN protocol

Interpretation, and Network. This protocol mirrors three primary steps aligned with the GNN model:

### Recommendation

Leveraging embedding vectors generated by GNN, suggest pancreatic cancer drug combinations by identifying those with the highest similarity or synergy scores.

### Analysis

Employing natural language processing, scour relevant articles from clinical trials involving the recommended drugs, extracting pertinent details such as dosage, outcomes, and side effects.

### Interpretation and Network meta-analysis

Employing network meta-analysis, evaluate the effectiveness of recommended drug combinations based on clinical trial data, and rank them accordingly. Additionally, elucidate the underlying mechanisms of drugs and their interactions with genes or proteins associated with pancreatic cancer.

Like several recently published medical AI papers, the RAIN protocol stands out by utilizing artificial intelligence to address a particular medical question. [215], [216]

#### 2.1. Stage I: **Recommendation**

A Graph Attention Network (GAT) represents a neural network designed to learn from graph-structured data, such as networks involving drugs and proteins. Utilizing a mechanism known as *attention*, GAT assigns varying weights to neighboring node features within a graph, depending on their relevance to the node under consideration. This adaptive approach enables GAT to grasp the intricate and diverse relationships among graph nodes without necessitating predefined graph operations or prior knowledge of the graph structure.

In the context of our subject, our GAT was deployed to propose drug combinations for pancreatic cancer by drawing insights from a knowledge graph containing diverse biomedical data, including drug-protein interactions, gene expression, and drug-target interactions. Through attention-based feature analysis of graph nodes, GAT generated *embedding vectors* for each drug and protein. These numerical representations encapsulate their characteristics and associations, facilitating measurement of similarity or synergy between different drugs and ranking of potential drug combinations for the disease. GAT boasts several noteworthy features, including its utilization of a multi-head attention mechanism. This mechanism enables

GAT to discern various types of attention weights for each node within the graph. Consequently, GAT can grasp diverse facets of node features and relationships, amalgamating them to form a more intricate and resilient embedding vector.

Furthermore, GAT demonstrates proficiency in handling heterogeneous graphs, which encompass nodes and edges of differing types.

Within the subject’s knowledge graph, nodes represent drugs and proteins, while edges signify diverse interactions like drug-protein, drug-target, and gene expression. GAT adeptly learns distinct attention weights for each edge type, allowing for tailored aggregation of neighboring node features. This enables GAT to extract pertinent insights from the varied and intricate information within the heterogeneous graph, thus producing embedding vectors that are both relevant and precise.

Moreover, GAT exhibits the capability to be seamlessly stacked into multiple layers, thereby enabling learning from higher-order neighborhoods within the graph. Within the subject’s knowledge graph, a drug node may have direct neighbors consisting of protein nodes and indirect neighbors comprising other drug nodes. Through the stacking of multiple GAT layers, the drug node can assimilate information from both its direct and indirect neighbors, enriching its embedding vector with their features. This holistic approach allows GAT to glean insights from the global structure of the graph, culminating in more comprehensive and insightful embedding vectors.

These distinguishing features position GAT as a potent and adaptable model for leveraging graph-structured data, particularly in suggesting drug combinations for pancreatic cancer.

Through the integration of GAT within the RAIN protocol, we unveil a novel and efficacious drug combination for pancreatic cancer, comprising Gemcitabine, Pancrelipase Amylase, and Octreotide. Our approach holds promise in guiding healthcare professionals and researchers toward optimal patient treatments while enhancing our understanding of the disease

#### 2.2. Stage II: **Analysis** - by comprehensive Systematic Review

In this phase, we outline a method for validating the outcomes of a GNN model by systematically evaluating suggested medications. We conduct a thorough systematic review utilizing databases including Science Direct, Embase, Scopus, PubMed, and Web of Science, alongside Google Scholar, to gather previously published articles for assessment.

Rather than conducting manual searches through databases, we utilize a semantic search approach powered by Natural Language Processing (NLP). This approach conducts individual searches for each term within MeSH, allowing for the discovery of a broader and more accurate selection of articles within a relatively short period.

### Information sources

We have employed an NLP-driven systematic review to locate pertinent studies across multiple databases, encompassing Science Direct, Embase, Scopus, PubMed, Web of Science, and Google Scholar. The aim is to verify the suggested medication combination created by an internal GNN model, through the analysis of data extracted from a sizable clinical trial. Keywords are derived from the outcomes of the GNN model and the Malignant neoplasm of pancreas subscription.

### Search strategy

A semantic search is performed using natural language processing (NLP) to explore titles and abstracts of publications across diverse databases. This approach enables the inclusion of MeSH terms as potential search terms, capitalizing on the benefits of semantic search.

### Study selection

Initially, duplicate studies are eliminated from the outset of the process. Following this, a thorough list encompassing all remaining research titles is assembled during the assessment phase to aid in organized filtering of the research materials. In the initial phase of the systematic review, referred to as screening, the titles and abstracts of the remaining research are carefully examined, and particular studies are omitted based on predetermined selection criteria.

In the second phase, known as competency assessment, the full texts of the research articles identified during the screening phase are thoroughly examined according to predefined selection criteria, leading to the exclusion of numerous irrelevant studies. In order to reduce the impact of individual bias on the selection of resources, both an expert and an NLP Question-Answering (QA) agent independently conduct research and data extraction. The expert is required to furnish a comprehensive rationale for any study that isn’t selected for inclusion. The quality assurance agent evaluates each article by assigning a score based on predefined questions, and articles with the least favorable ratings are omitted. These inquiries focus on the effectiveness of medications in treating Malignant neoplasm of pancreas, with different drugs substituted for “this drug” in each query generated by the intelligent system. If there is disagreement between the evaluations of the expert and the QA agent, the expert will reassess the disputed research.

### Quality evaluation

A checklist is employed to assess the quality of the remaining publications, tailored to the particular type of research being undertaken. Typically, The STROBE method is used to assess the quality of observational studies. This checklist comprises six primary sections: title, abstract, introduction, methodology, results, and discussion, covering a total of 32 fields, which also include subcategories.

Each of the 32 aspects listed in the checklist pertains to a specific facet of the study methodology, encompassing components like the title, issue description, research goals, study type, target population, sampling method, sample size, variable definitions and procedures, data collection techniques, and statistical analysis, approaches, and outcomes. A maximum score of 32 is achievable in the quality assessment conducted with the STROBE checklist. Articles that score 16 or higher are categorized as demonstrating moderate to high quality.

#### 2.3. Stage III: **Interpretation** and **Network** meta-analysis

During the third stage, a network meta-analysis is utilized to assess the influence of recommended artificial drug combinations on human proteins/genes. This entails analyzing multiple drugs simultaneously within a single study. This approach merges both direct and indirect data regarding the relationship between the disease and drugs, utilizing proteins/genes as a connecting element within a network of randomized controlled trials. It assists in assessing the relative effectiveness of frequently prescribed medications in clinical settings. The effectiveness of each medication is gauged using biological data as input.

## 3. RESULTS

### 3.1. Stage I: **Recommendation** – using GAT

The drug combination suggested by the GNN comprises Gemcitabine, Amylase, and Octreotide. Table 1 presents the p-values linked to the amalgamation of these drugs. For example, the p-value related to pancreatic malignant neoplasms and Gemcitabine (Scenario 1) is 0.018113539, decreasing to 5.40719E-05 with the addition of Amylase (Scenario 2). The p-value resulting from the implementation of the third scenario, 3.94E-07, indicates that the proposed drug combination positively influenced disease management. Table 2 illustrates the alterations in p-values between human proteins/genes and Malignant neoplasm of pancreas with different scenarios. The ‘S0’ column displays the p-value between Malignant neoplasm of pancreas and the respective influenced human proteins/genes. The ‘S1’ column indicates the collective p-value when Gemcitabine is utilized. In the ‘S3’ column, numerous p-values between Malignant neoplasm of pancreas and human proteins/genes approach 1, suggesting a diminishing significance of the target proteins/genes.

**Table 1:** p-value between scenarios and Malignant neoplasm of pancreas.

| *Scenario* | *drug combination* | *p-value* |
| --- | --- | --- |
| S1 | Gemcitabine | 0.018113539 |
| S2 | S1+Amylase | 5.40719E-05 |
| S3 | S2+Octreotide | 3.94048E-07 |

**Table 2:** p-values between Malignant neoplasm of pancreas and human peroteins and genes after implementing scenarios.

| Association Name | S0 | S1 | S2 | S3 | Association Name | S0 | S1 | S2 | S3 |
| --- | --- | --- | --- | --- | --- | --- | --- | --- | --- |
| KRAS | 1.24E-05 | 0.999784 | 0.999784 | 0.999784 | FOXM1 | 0.000307 | 0.988543 | 0.988543 | 0.988543 |
| SST | 2.39E-05 | 2.39E-05 | 0.999727 | 1 | MUC2 | 0.000308 | 0.69604 | 0.990078 | 0.990078 |
| MEN1 | 2.51E-05 | 2.51E-05 | 2.51E-05 | 0.999531 | RNF43 | 0.000309 | 0.780996 | 0.981126 | 0.981126 |
| SMAD4 | 2.82E-05 | 0.997365 | 0.997365 | 0.997497 | VIM | 0.000312 | 0.997451 | 0.999977 | 0.999977 |
| CHGA | 7.31E-05 | 7.31E-05 | 0.997639 | 1 | PARP1 | 0.000314 | 0.999602 | 0.999711 | 0.999711 |
| GAST | 9.2E-05 | 0.13289 | 0.999667 | 1 | GNAS | 0.000322 | 0.846152 | 0.991375 | 0.99994 |
| MSLN | 9.6E-05 | 0.995936 | 0.997036 | 0.997036 | S100P | 0.000326 | 0.988341 | 0.988341 | 0.988341 |
| LOC100508689 | 0.000102 | 0.98717 | 0.999991 | 0.999991 | COL11A2 | 0.000328 | 0.999566 | 0.999566 | 0.999566 |
| PALB2 | 0.000103 | 0.985191 | 0.985191 | 0.985191 | DAXX | 0.000338 | 0.000338 | 0.000338 | 0.965823 |
| CDKN2A | 0.000107 | 0.990081 | 0.999052 | 0.999687 | GLI1 | 0.000363 | 0.996384 | 0.996384 | 0.996384 |
| BRCA2 | 0.000108 | 0.997677 | 0.997677 | 0.998743 | MUC5AC | 0.000366 | 0.958025 | 0.996718 | 0.996718 |
| TP53 | 0.000108 | 0.999518 | 0.999983 | 0.999993 | MUC6 | 0.000371 | 0.000371 | 0.979142 | 0.979142 |
| MUC1 | 0.00011 | 0.997683 | 0.999894 | 0.999894 | PDX1 | 0.000382 | 0.95897 | 0.999943 | 0.999943 |
| PPY | 0.000126 | 0.000126 | 0.999721 | 1 | SYP | 0.000383 | 0.125144 | 0.988445 | 0.999929 |
| MUC4 | 0.000139 | 0.998155 | 0.999499 | 0.999499 | BRCA1 | 0.000395 | 0.997745 | 0.997745 | 0.99931 |
| PRSS1 | 0.000162 | 0.000162 | 0.999522 | 0.999522 | BIRC5 | 0.000397 | 0.999413 | 0.999899 | 0.999899 |
| GCG | 0.000167 | 0.892831 | 0.999958 | 1 | ZEB1 | 0.000406 | 0.99816 | 0.99816 | 0.99816 |
| PRSS58 | 0.000168 | 0.695651 | 0.998671 | 0.998671 | EPCAM | 0.000413 | 0.984197 | 0.984197 | 0.984197 |
| SLC29A1 | 0.00017 | 0.999987 | 0.999987 | 0.999987 | MTOR | 0.000437 | 0.998758 | 0.999885 | 1 |
| CDH1 | 0.000173 | 0.997992 | 0.997992 | 0.997992 | STAT3 | 0.000465 | 0.997966 | 0.999201 | 0.999201 |
| SCT | 0.00018 | 0.00018 | 0.999989 | 1 | CCND1 | 0.000477 | 0.997585 | 0.999404 | 0.999464 |
| SSTR2 | 0.000189 | 0.789838 | 0.789838 | 1 | CELA3B | 0.000478 | 0.000478 | 0.999876 | 0.999876 |
| TSC1 | 0.000194 | 0.99515 | 0.999807 | 0.999807 | EGFR | 0.000482 | 0.999791 | 0.999993 | 0.999993 |
| SPINK1 | 0.00029 | 0.89844 | 0.999964 | 0.999969 | DCK | 0.000484 | 0.999994 | 0.999994 | 0.999994 |
| CELA3A | 0.000302 | 0.000302 | 0.999924 | 0.999924 | PSCA | 0.000527 | 0.524185 | 0.524185 | 0.524185 |

### 3.2. Stage II: A comprehensive Systematic Review

This phase investigates the impact of the mentioned drugs on Malignant neoplasm of pancreas treatment. Articles focusing on this aspect were collected and methodically evaluated, following PRISMA guidelines and the RAIN framework. Initially, 309 possibly pertinent articles were recognized and transferred into the EndNote citation management system. Among them, 175 duplicates were removed. Subsequently, 134 papers underwent screening based on their titles and abstracts, resulting in the exclusion of 50 studies. Eligibility evaluation reduced the pool to 84 studies, with 53 studies being excluded after full-text review and consideration of inclusion/exclusion criteria. In the quality assessment stage, 10 out of the remaining 31 studies were eliminated due to low scores on the STROBE checklist and methodological shortcomings, leaving 21 cross-sectional studies for final analysis. The complete texts of the articles underwent analysis, with each paper evaluated using the STROBE checklist, depicted in Figure 3. The chemical structures of the drugs are illustrated in Figure 4, while Table 3 presents the characteristics of the drugs. Comprehensive details and attributes of these articles are outlined in Table 4. [217], [218], [219], [220], [221], [221], [222], [223], [224], [225], [226], [227], [228], [229], [230], [231], [232], [233], [234], [235], [236], [237]

**Table 3:** Characteristics of suggested medications as efficacious treatments for managing pancreatic malignant neoplasms.

| Drug Name | Accession Number | Type |  | Formula | Mechanism of Action |
| --- | --- | --- | --- | --- | --- |
|  |  | Small Molecule | Biotech |  |  |
| Gemcitabine | DB00441 | * | | $C_9H_{11}F_2N_3O_4$ | Gemcitabine, a potent deoxycytidine analog, is phosphorylated within cancer cells to create active forms dFdCDP and dFdCTP. These metabolites interrupt DNA elongation, inducing cell death through chain termination. Additionally, gemcitabine inhibits ribonucleotide reductase, reducing dCTP levels and facilitating the integration of dFdCTP into DNA. This mechanism enhances gemcitabine's therapeutic efficacy and sustains elevated levels of active metabolites inside cells, setting it apart from cytarabine. |
| Amylase | DB11065 | | * | $(C_6H_{12}O_6)_x$ | Pancrelipase amylase functions by compensating for the deficiency of natural amylase in the body. It works by breaking down alpha 1-4 linkages in polysaccharides containing three or more glucose units through hydrolysis. However, it does not hydrolyze alpha 1-6 linkages, leaving starch partially reduced to smaller molecules. |
| Octreotide | DB00104 | | * | $C_{49}H_{66}N_{10}O_{10}S_2$ | Octreotide binds to somatostatin receptors, leading to smooth muscle contraction in blood vessels. This process involves activating phospholipase C and inhibiting L-type calcium channels, resulting in decreased growth hormone levels. These actions effectively treat the various growth hormone-related and metabolic effects seen in acromegaly. |

**Table 4:** some important research studies for proposed drugs in Malignant neoplasm of pancreas managements.

| **study name** | **Gencitabine** | **AMYLASE** | **Octreotide** |
| --- | --- | --- | --- |
| Gailhouste L. et al., 2018 | * |  | * |
| Zhang G. et al., 2016 |  | * | * |
| Kapoor VK. et al., 2016 |  | * | * |
| Suleiman Y. et al., 2015 | * |  | * |
| Maegawa Y. et al., 2014 | * | * |  |
| Anders M. et al., 2014 |  | * | * |
| Saif MW. et al., 2011 | * |  | * |
| Sui C. et al., 2009 | * |  | * |
| Mouri H. et al., 2008 | * |  | * |
| Nayak TK. et al., 2008 | * |  | * |
| Closset J. et al., 2008 |  | * | * |
| Antoine M. et al., 2007 | * |  | * |
| Dauendorffer JN. et al., 2007 |  | * | * |
| Thomopoulos KC. et al., 2006 |  | * | * |
| Hadjicostas P. et al., 2006 |  | * | * |
| Polyzos A. et al., 2005 | * |  | * |
| Kouloulis VE. et al., 2002 |  | * | * |
| Emoto T. et al., 1997 |  | * | * |
| Schnall SF. et al., 1996 | * |  | * |
| Durden FM. et al., 1996 |  | * | * |
| Bianchi G. et al., 1993 |  | * | * |

**Figure 3:**
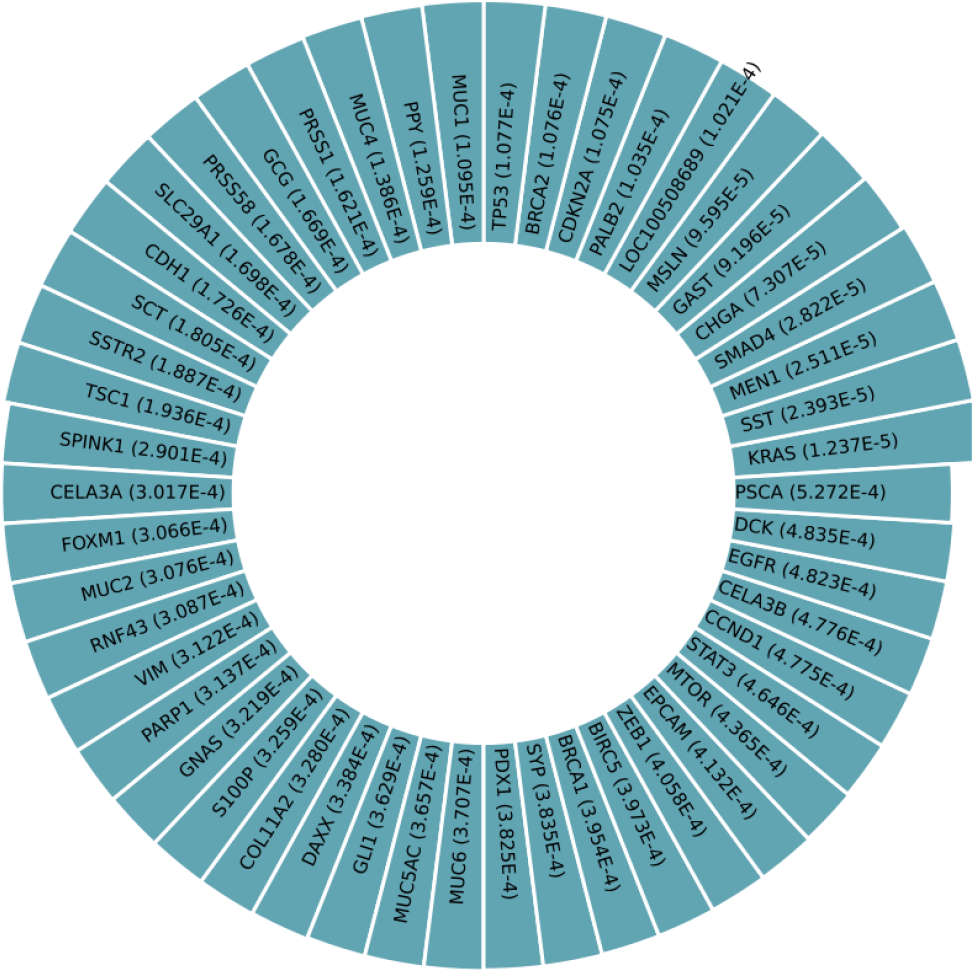
p-values between affected human proteins/genes and Malignant neoplasm of pancreas

**Figure 1:**
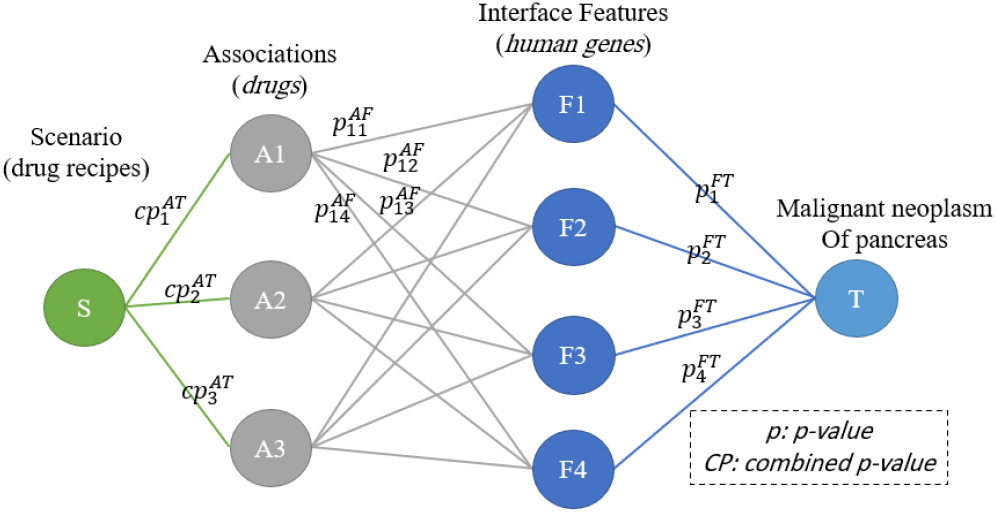
The general structure of the GNN model to suggest an effective drug combination in the management of disease using human proteins/genes as interface features

### 3.1. Stage III: Interpretation and Network meta-analysis

Figure 5 showcases the p-values. associated with human proteins/genes impacted by Malignant neoplasm of pancreas, whereas Figure 6 illustrates the p-values subsequent to the implementation of the third scenario. Figure 7 employs a radar chart to demonstrate the efficacy of drugs identified by the drug selection algorithm, showcasing the p-values between Malignant neoplasm of pancreas and human proteins/genes after the application of the chosen medications. Figure 8 illustrates the p-values indicating associations between targets and various interface features. Green and blue colors represent p-values below .01 and .05, respectively. Each line of different colors corresponds to the efficacy of the specific drug in that particular situation.

**Figure 2:**
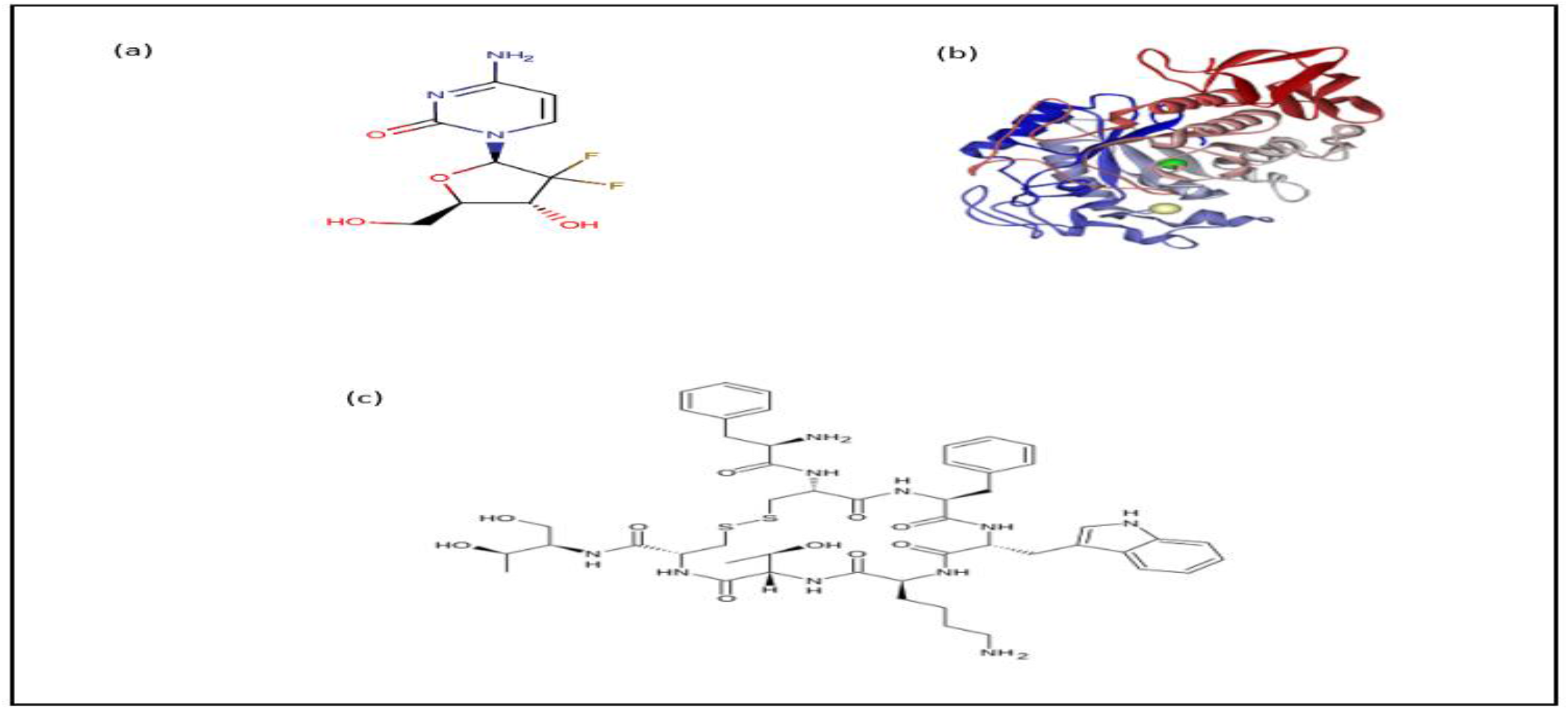
Drug structure for (a) Gemcitabine, (b) Amylase, (c) Octreotide from https://www.drugbank.com/

**Figure 6:**
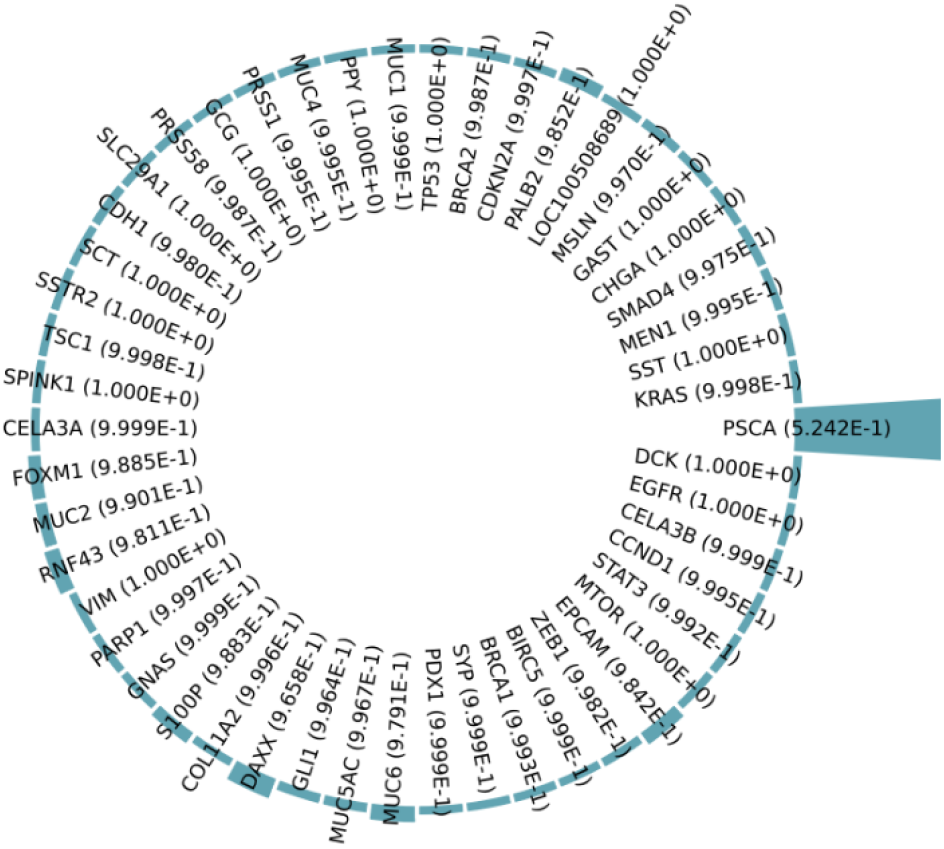
p-values between affected human proteins/genes and Malignant neoplasm of pancreas after implementing Scenario 4.

**Figure 7:**
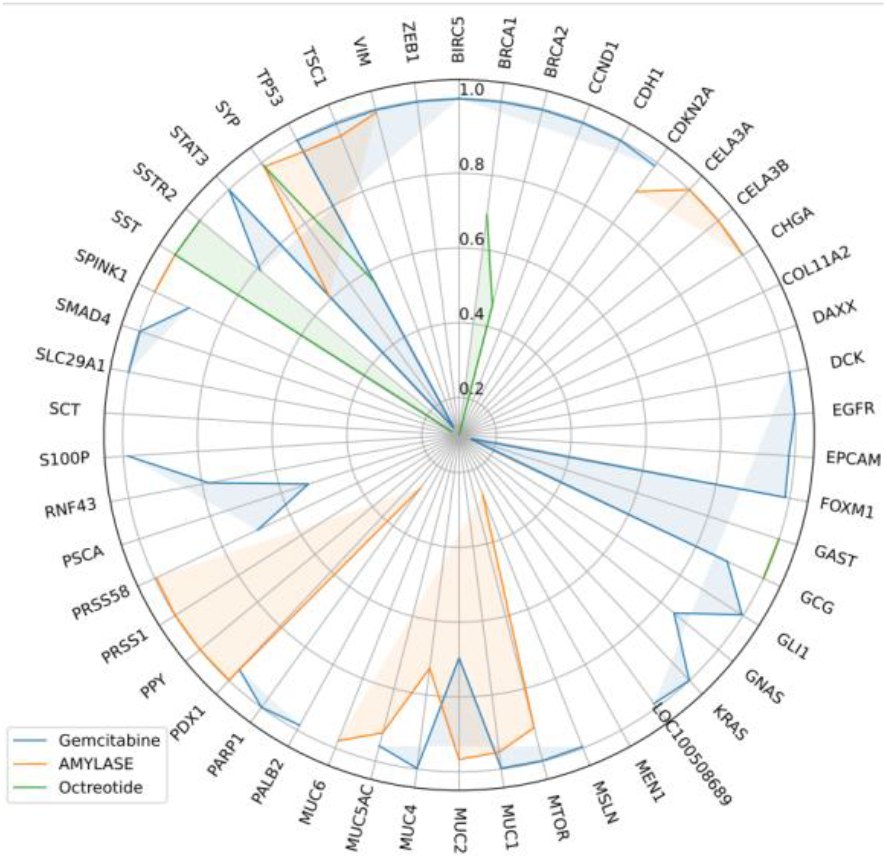
radar chart for p-values between Malignant neoplasm of pancreas and affected proteins/genes, after consumption of each drug

**Figure 8:**
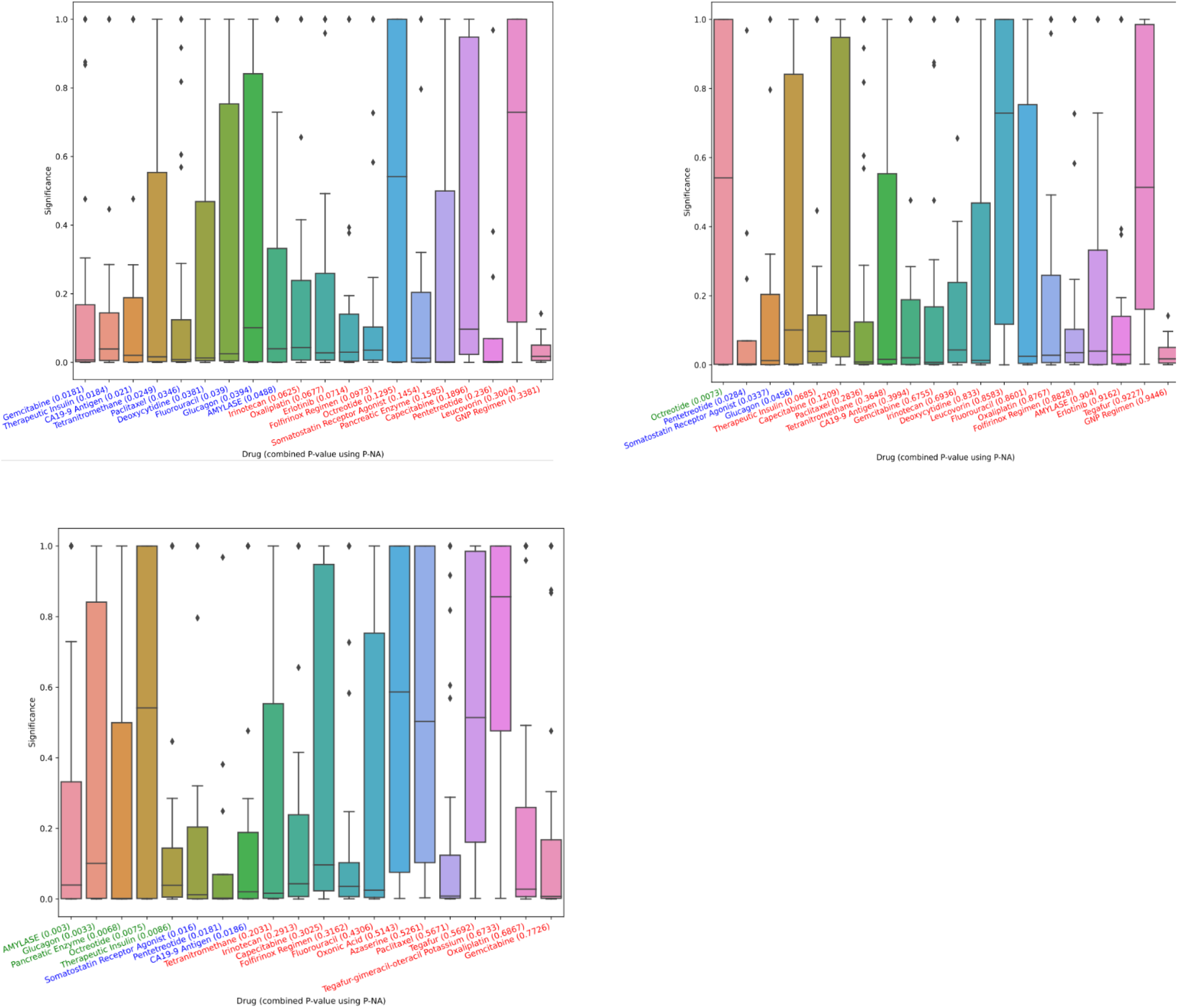
p-values between associations and target, using different interface features. (a) Overall, (b) after first drug from is used, (c) after first drugs of (a) and (b) are used.

## 4. DISCUSSION

Information concerning prescribed medications is examined to explore possible interactions with other drugs, compatibility with food, adverse reactions, and considerations for serious health conditions. Reputable online platforms such as Medscape, WebMD, Drugs, and Drug bank are consulted to compare medications directly. These databases scrutinize pairs of drugs, uncovering potential interactions among specific combinations. Currently, none of these platforms indicate significant interactions between Gemcitabine, Amylase and Octreotide.

Gemcitabine is a chemotherapy medication commonly used to treat various types of cancer, including pancreatic cancer, breast cancer, lung cancer, and ovarian cancer. Amylase, on the other hand, is an enzyme produced by the pancreas and salivary glands that helps in the digestion of carbohydrates. While there isn’t a direct drug interaction between gemcitabine and amylase, it’s important to understand their respective effects and potential side effects. Gemcitabine is primarily metabolized by the liver and excreted through the kidneys. It can cause various side effects, including myelosuppression (reduced bone marrow activity leading to low blood cell counts), nausea, vomiting, fatigue, and flu-like symptoms. Amylase levels may be monitored in cancer patients because elevated levels of serum amylase can be indicative of pancreatic inflammation or damage, which can be a side effect of gemcitabine therapy. Pancreatitis is a known adverse effect associated with gemcitabine treatment, albeit it’s relatively rare. If a patient experiences symptoms such as severe abdominal pain, nausea, vomiting, or fever during gemcitabine treatment, it’s essential to promptly evaluate them for potential pancreatitis.

Amylase and octreotide are two different drugs with distinct mechanisms of action and therapeutic uses. Amylase and octreotide serve different purposes and have distinct mechanisms of action, there are no known direct drug interactions between them.

Gemcitabine and octreotide are two medications that are used for different purposes, but they can potentially interact with each other. Gemcitabine is a chemotherapy medication used to treat various types of cancer, including pancreatic cancer. Octreotide is a medication primarily used to treat conditions such as acromegaly, carcinoid syndrome, and certain types of tumors. There isn’t a well-documented direct drug interaction between gemcitabine and octreotide, but it’s essential to note that both medications can affect the function of the pancreas. Octreotide can reduce the secretion of insulin and other pancreatic hormones, while gemcitabine may cause pancreatic inflammation or pancreatitis as a side effect. Therefore, using these medications together might potentially increase the risk of pancreatic complications. Additionally, both medications can have effects on blood cell counts. Gemcitabine may cause bone marrow suppression, leading to low blood cell counts (neutropenia, thrombocytopenia, anemia), while octreotide may rarely cause changes in blood sugar levels and gallbladder function.

When these three medications are used together, there is a potential for interactions, especially concerning pancreatic function. Gemcitabine may exacerbate pancreatic issues, while octreotide can further suppress pancreatic hormone secretion. Monitoring for signs of pancreatic complications, changes in blood cell counts, and alterations in blood sugar levels is essential when using this combination of medications.

## 5. CONCLUSION

Pancreatic cancer presents significant challenges necessitating innovative therapeutic approaches. In this investigation, we introduced a novel methodology merging graph attention networks (GATs) with the RAIN protocol to identify optimal drug combinations for targeting the genes and proteins associated with pancreatic cancer. We illustrated the capability of our approach in recommending a synergistic drug trio— Gemcitabine, Pancrelipase Amylase, and Octreotide— that exhibits favorable effects on disease outcomes. Our findings were substantiated through analysis of clinical trials and literature, alongside network meta-analysis comparing the efficacy of our drug combination against existing treatments. This method represents a robust tool for both drug discovery and elucidating disease mechanisms, with potential applicability to other intricate diseases.

## Data Availability

Datasets are available through the corresponding author upon reasonable request.

## Abbreviations

STROBE: Strengthening the Reporting of Observational studies in Epidemiology; PRISMA: Preferred Reporting Items for Systematic Reviews and Meta-Analysis; RAIN: Systematic Review and Artificial Intelligence Network Meta-Analysis

## Acknowledgements

By researchers in Bioinformatics, Computational Biology, Artificial Intelligence and Medicine.

## Authors’ contributions

EP contributed to design, AAK and MB native RL algorithm, EP, AAK, MB bio-statistical analysis; MB participated in most of the study steps. AAK used GAT in exploring drug combinations. All authors have read and approved the content of the manuscript

## Availability of data and materials

Datasets are available through the corresponding author upon reasonable request.

## Ethics approval and consent to participate

Not applicable.

## Consent for publication

Not applicable.

